# Laminar multi-contrast fMRI at 7T allows differentiation of neuronal excitation and inhibition underlying positive and negative BOLD responses

**DOI:** 10.1101/2024.04.01.24305167

**Authors:** Xingfeng Shao, Fanhua Guo, JungHwan Kim, David Ress, Chenyang Zhao, Qinyang Shou, Kay Jann, Danny JJ Wang

## Abstract

A major challenge for human neuroimaging using functional MRI is the differentiation of neuronal excitation and inhibition which may induce positive and negative BOLD responses. Here we present an innovative multi-contrast laminar functional MRI technique that offers comprehensive and quantitative imaging of neurovascular (CBF, CBV, BOLD) and metabolic (CMRO_2_) responses across cortical layers at 7 Tesla. This technique was first validated through a finger-tapping experiment, revealing ’double-peak’ laminar activation patterns within the primary motor cortex. By employing a ring-shaped visual stimulus that elicited positive and negative BOLD responses, we further observed distinct neurovascular and metabolic responses across cortical layers and eccentricities in the primary visual cortex. This suggests potential feedback inhibition of neuronal activities in both superficial and deep cortical layers underlying the negative BOLD signals in the fovea, and also illustrates the neuronal activities in visual areas adjacent to the activated eccentricities.

## 1. Introduction

High resolution functional magnetic resonance imaging (fMRI) at ultrahigh field has significantly enhanced our capability to study neurovascular and metabolic activities in-vivo at the mesoscopic scale. A major challenge for fMRI, however, is the differentiation of neuronal excitation and inhibition which may induce positive and negative BOLD responses, depending on the complex interplay between cerebral blood flow (CBF), cerebral blood volume (CBV), and cerebral metabolic rate of oxygen (CMRO_2_) (Harel et al., 2002; Huber et al., 2019; Shmuel et al., 2002). The relationship between changes in neuronal activities and changes in fMRI signals has long been a focus of debate, especially regarding negative BOLD responses (NBR). Shmuel et al. hypothesized two possibilities for the formation of NBR: 1) from the reduction of neuronal activity; and 2) from hemodynamic changes independent of neuronal activity (Shmuel et al., 2002). NBR has been shown to be associated with reduction in CBF, CBV and neuronal activity (Boorman et al., 2010; Devor et al., 2007; Harel et al., 2002; Northoff et al., 2007; Shmuel et al., 2006). However, other studies have found that NBR may occur with heightened neuronal activity (Goense et al., 2012; Mullinger et al., 2014; Schridde et al., 2008). These contradicting findings highlight the complexity of the NBR and BOLD signals. As illustrated in Figure 1, different scenarios for changes in CBF, CBV and CMRO_2_ could lead to positive or negative BOLD responses respectively. This is a major challenge to precisely interpret the underlying neural mechanisms using existing laminar fMRI techniques that measure a single hemodynamic parameter (Fracasso et al., 2018; Han et al., 2021; Huber et al., 2017; Shao et al., 2021).

**Figure 1.**
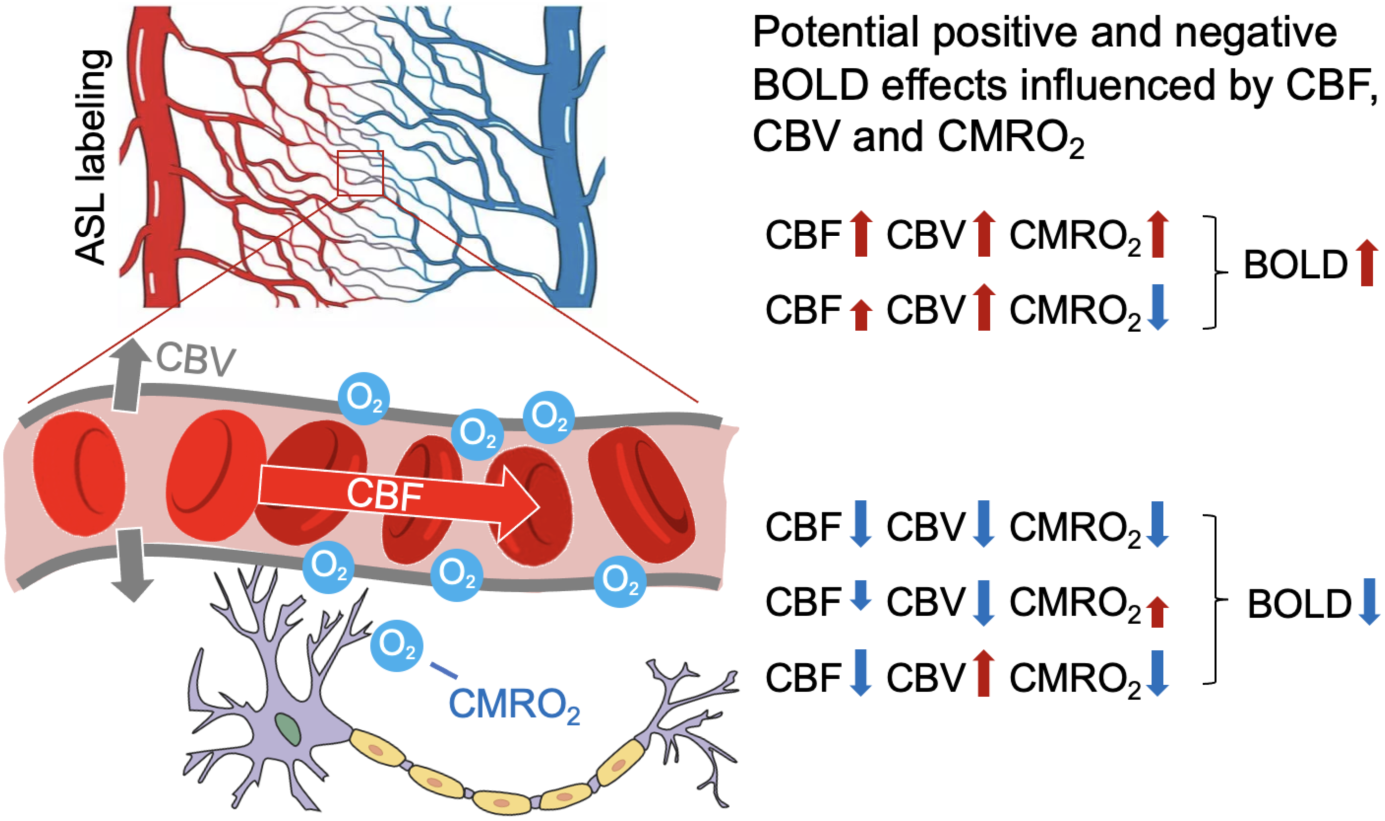
Illustration of neurovascular responses induced by neuronal activities. The diagram demonstrates the relationship between neuronal activation and its resultant increase in oxygen consumption, accompanied by both cerebral blood flow (CBF) and volume (CBV) increase. This increased concentration of oxygenated hemoglobin leads to the generation of positive BOLD signals, as commonly seen in fMRI studies. Given the complex dynamics between CBF, CBV, and CMRO_2_ during various neuronal activities, BOLD signals may present as either positive and negative, and potential mechanisms contributing to positive and negative BOLD signals are shown in the middle.

In vivo imaging of the full set of hemodynamic parameters including CBF, CBV and BOLD as well as metabolic parameters such as CMRO_2_ provides comprehensive characterization of neurovascular responses, and precise inference of the neuronal activities. In particular, changes in CMRO_2_ are closely related to changes in neuronal activity measured by electrophysiology across cortical layers (Herman et al., 2013). Yang et al first proposed a technique at 3 Tesla to simultaneously measure CBF, CBV and BOLD contrasts by combining pulsed arterial spin labeling (ASL) with vascular space occupancy (VASO) (Yang et al., 2004), which was further developed by Cheng et al (Cheng et al., 2017) at 3T and adapted for 7T by Krieger et al (Krieger et al., 2015). However, pulsed ASL is susceptible to vascular contaminations due to arterial transit effects (Shao et al., 2019). In addition, the spatial resolution and/or image coverage of the above techniques does not allow laminar fMRI. Optical imaging in animals provides evidence that CBF can be precisely regulated on an extremely fine scale at the level of capillaries (Chaigneau et al., 2003; Hall et al., 2014; Hamilton et al., 2010; Peppiatt et al., 2006). Therefore, it is highly desirable to develop high resolution multi-contrast fMRI at ultrahigh field for simultaneous hemodynamic and metabolic imaging at laminar level.

Here we present an innovative multi-contrast laminar fMRI technique at 7 Tesla, which enables simultaneous measurement of CBF, CBV, T2-weighted BOLD, and CMRO_2_. The key innovation of the proposed method lies in the use of pseudo-continuous ASL (pCASL) with intracranial labeling and a zoomed 3D gradient-and-spin echo (GRASE) readout, coupled with a non-selective inversion pulse at an optimized post-labeling delay (PLD), to generate concurrent ASL perfusion and VASO contrast for CBV. T2-weighted BOLD signals, which has improved specificity than T2*-weighted BOLD, were acquired after the ASL/VASO acquisition with GRASE readout. Compared to existing laminar fMRI methods, our technique offers several unique advantages: 1) CBF, CBV, T2-weighted BOLD contrasts are simultaneously measured to enable detailed and precise characterization of hemodynamic changes across cortical layers at 7T; 2) Concurrent CBF, CBV, T2-weighted BOLD contrasts enable accurate estimation of the laminar profile of CMRO_2_ using calibrated fMRI with a respiratory challenge (breath-hold) and an innovative algorithm to estimate laminar profiles of calibration parameters; 3) The use of pCASL with optimized PLD offers higher SNR than pulsed ASL, while minimizing pial arterial signals to ensure that the measured perfusion signal arises from capillaries and brain tissue (Shao et al., 2021).

In this study, we performed 4 experiments to demonstrate the capability of the proposed multi-contrast fMRI for comprehensive neurovascular and metabolic imaging to understand the underlying neuronal mechanisms associated with positive and negative BOLD responses: 1) Validating the specificity of our proposed technique by measuring activations in both deep and superficial layers of the primary motor cortex (M1) induced by finger-tapping (FT) tasks; 2) Estimating calibrated fMRI parameters through a breath-hold (BH) hypercapnia method, enabling the calculation of CMRO_2_ (Davis et al., 1998) from simultaneously measured CBF, CBV, and BOLD signals; 3) Conducting eccentricity mapping using the population receptive field (pRF) approach (Greene et al., 2014), a technique that quantifies visual field representations in the brain to understand visual processing and cortical organization; 4) Performing a ring-shaped visual stimulation that elicited positive and negative BOLD responses at the projected visual area and the fovea respectively, while investigating layer-dependent CBF, CBV, T2-weighted BOLD, and CMRO_2_ activities and their variations across eccentricities, aiming to understand the underlying neuronal activities associated with positive and negative BOLD responses.

## 2. Methods

### 2.1. MRI pulse sequence

The diagram of MRI pulse sequence is shown in Fig. 2A. Perfusion contrast was generated by pCASL, and a non-selective *hyperbolic secant (HS*)-inversion pulse was added after pCASL pulse train to suppress background signal and generate VASO contrast (Shao et al., 2021). Due to limited B1+ at bottom of brain at 7T, we assume fresh in-flow blood only experience one inversion and control images acquired at PLD = 1160 msec, which nulls in-flow blood signal (T_1,blood_ = 2.1sec (Rane & Gore, 2013)), exhibit VASO contrast. The PLD = 1160 msec is slightly longer than ATT in M1 and V1, which provides desirable SNR for functional ASL study (Shao et al., 2021). Concurrent T2-BOLD signals were acquired 500 msec after the ASL/VASO acquisition (Fig. 2A). Intracranial vasculature was revealed by maximal-intensity-projection (MIP) of the MP2RAGE (0.7mm^3^) images, and the pCASL labeling plane was placed above the circle of Willis (CoW) and simultaneously perpendicular to the anterior cerebral artery (ACA), middle cerebral artery (MCA) and posterior cerebral artery (PCA) (Fig. 2B). This labeling location took advantage of the higher B1 and more homogeneous B0 field around the center of the brain. Imaging parameters were: FOV = 80×40 mm^2^, 14 slices, 1-mm^2^ in-plane resolution, 2.2 mm slice thickness, 3 segments along phase direction, TE = 17.2 msec, labeling duration = 1280 msec, TR = 4000 msec. Volume TR was 24 sec for one CBF and VASO image, and 12 sec for one BOLD image (Fig. 2C).

**Figure 2.**
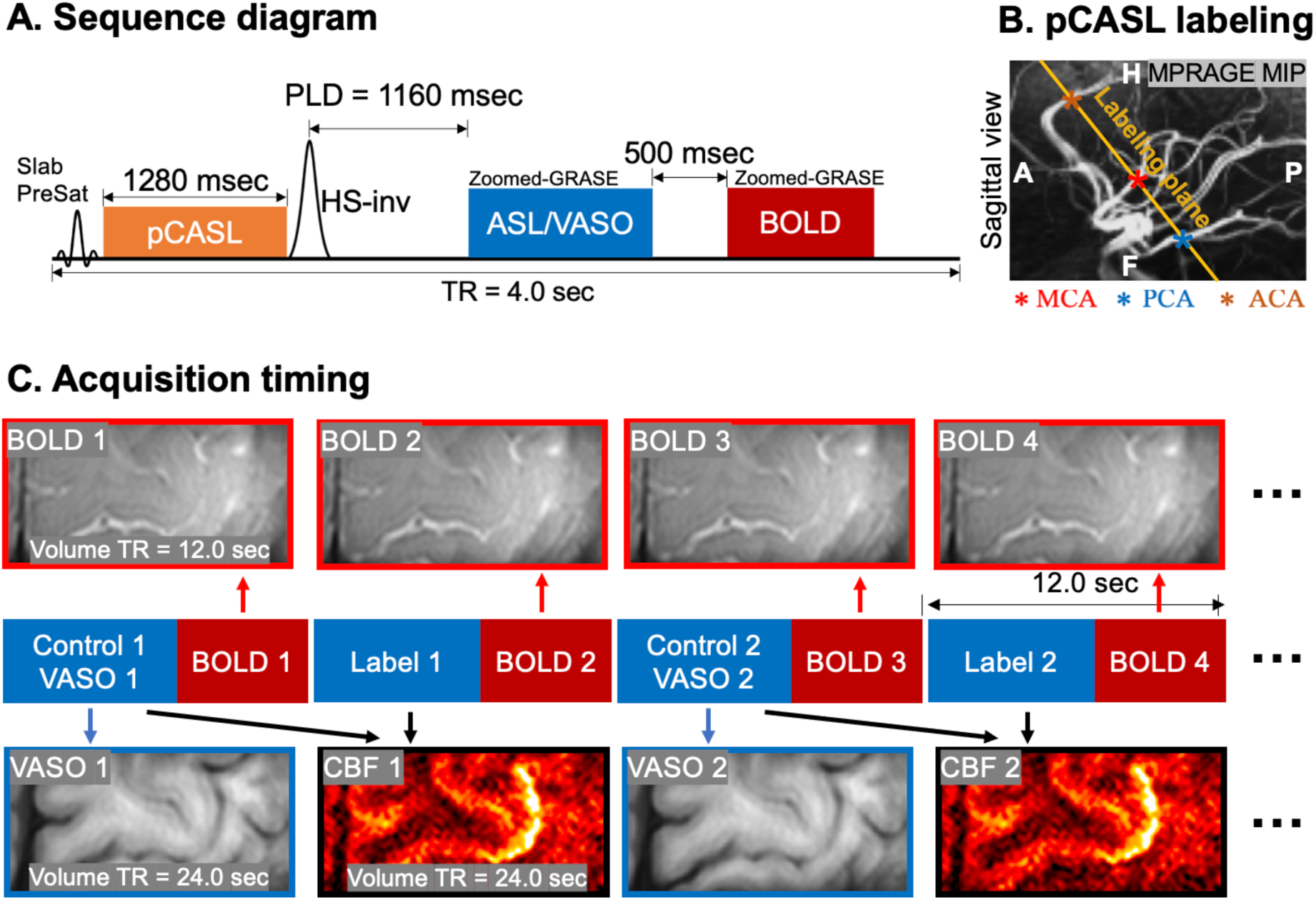
(A) MRI pulse sequence diagram. A non-selective hyperbolic secant (HS)-inversion pulse was added after pCASL labeling train to suppress background signal and generate VASO contrast (ASL control). PLD = 1160 msec to null fresh in-flow blood signals. BOLD signal was acquired 500 msec after ASL/VASO acquisition. TR = 4 sec for one segment, and volume TR = 12 sec for BOLD and 24 sec for ASL and VASO. (B) Position of the pCASL labeling plane, which was placed above the circle of Willis and perpendicular to MCA, PCA and ACA. (C) Acquisition timing. Each image volume was acquired in 3 segments (TR = 4 sec), so that volume TR for BOLD, CBF and VASO were 12, 24 and 24 sec, respectively.

### 2.2. MRI experiments

We conducted four experiments on a 7T Siemens Terra MRI scanner with 1Tx/32Rx head coil to validate the high sensitivity and specificity of the proposed technique in detecting neuronal activity and reveal the distinct neurovascular and metabolic responses across cortical layers and eccentricities under positive and negative BOLD signals induced by a ring-shape visual stimulus. Experiment details are shown as following:

#### 2.2.1. Experiment 1 – Finger tapping

Four runs with interleaved blocks of rest and sequential dominant-hand FT were acquired. Each run consisted of five blocks of interleaved rest and FT tasks (48 sec per block, 8 min each acquisition block). Total scan time was 32 min. As shown in Supplemental Fig. S1, we expect FT to induce neuronal activity in both superficial layers (hypothesize to involve sensory input from primary sensory cortex (S1) and premotor cortex) and deep layers (hypothesized to involve spinal output), and increased CBF and decreased VASO signals in both deep and superficial layers.

#### 2.2.2. Experiment 2 - Laminar profiles of M and β (Davis model) estimated by breath-hold (BH) induced hypercapnia

M and β are two unknown parameters which are likely to change across participants and cortical layers. Multi-contrast fMRI data were acquired with 12 cycles of interleaved 24 sec rest and 24 sec BH with visually presented instruction, with two runs collected in a total of 19 min 20 sec. Under the assumption that CMRO_2_ does not change during hypercapnia, we can directly estimate M and β using concurrently measured CBF, CBV, and BOLD signals between resting state and BH using the following simplified Davis model (Eq. [1]):

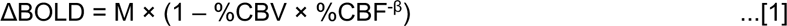

#### 2.2.3. Experiment 3 - Eccentricity mapping with a pRF stimulus

Eccentricity mapping was conducted through pRF analysis using a flickering checkerboard pattern that sweeps (30 steps, TR=1.2s) across the screen, sequentially stimulating different visual field regions with a 15-degree rotation after each sweep (Supplemental Fig. S3 top). We collected two pRF runs using 1.6 mm³ isotropic EPI (SMS-2, in-plane GRAPPA-2, TR=1.2s), and acquired 435 volumes in 8 min 42 sec for each run. Eccentricity masks ranging from 0 to 8 degrees within V1 were then reconstructed by estimating each voxel’s receptive field from the BOLD signal changes.

#### 2.2.4. Experiment 4 – Partial visual field activation stimulus

Finally, we conducted fMRI experiments using visual stimulus consisted of a mean-gray background with a ring-shaped sector subdivided into 12 subsectors, covering an eccentricity range of 4° to 6° (Supplemental Fig. S3 bottom). Each subsector contained a high-contrast (100%) radial grating (1 cycle per degree, cpd) that reversed contrast at a rate of 4Hz. Each run comprised six blocks of 48 sec of visual stimulus followed by 48 sec of rest, and four runs were acquired in a total of 39 minutes.

### 2.3. Human participants

All participants in this study were healthy without known neurological disorders and refrained from caffeine 5 hours before the scan. Experiment 1 included four participants (3 male, age = 25.3±3.2years). A total of nine participants were recruited for experiment 2 to 4 (3 male, age = 26.0±6.3 years). All participants provided written informed consents according to a protocol approved by the Institutional Review Board (IRB) of the University of Southern California. Head motion was minimized by placing cushions on top and two sides of head and taping participant’s chin to coil.

### 2.4. Data processing

#### 2.4.1. ASL data prep-processing

A separate proton density weighted M0 image with the same zoomed GRASE readout was acquired for ASL calibration. Dynamic ASL images were realigned and coregistered to M0 using SPM12 (Functional Imaging Laboratory, University College London, UK). Control and label images were pairwise subtracted to obtain raw perfusion images. Signal decay in k-space was prominent in partition direction due to long echo train length and short arterial blood T2 at 7T (68 msec (Krishnamurthy et al., 2014)). We implemented a singular value decomposition (SVD) based deblurring algorithm to reduce blurring along slice direction without amplifying noise (Shao et al., 2021). CBF was calculated from the relative perfusion signal (normalized by M0) using the one compartment model (Alsop et al., 2015; Buxton et al., 1998) with the following model parameters: Arterial blood T1 = 2.1 sec (Rane & Gore, 2013), tissue-to-blood partition coefficient λ = 0.9 mL/g (Alsop et al., 2015), labeling efficiency of 82.1% and 95% background suppression inversion efficiency (Shao et al., 2021). Task induced absolute CBF (ΔCBF) and relative CBF (%CBF) changes were obtained by subtraction (CBF_FT_ - CBF_rest_) and division (CBF_FT_ / CBF_rest_ -1) between task CBF and resting state CBF.

#### 2.4.2. VASO and BOLD data prep-processing

Task induced BOLD signal changes were calculated by:

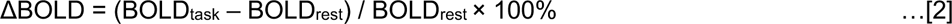

VASO signals were divided by BOLD to minimize BOLD effect and vein contamination (Huber, Ivanov, et al., 2014). Relative CBV (%) changes were calculated from VASO signal changes assuming baseline CBV_rest_ = 0.055 ml/ml (Lin et al., 2008; Lu et al., 2005):

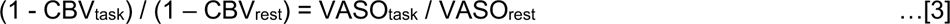

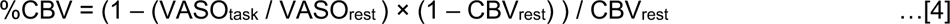

#### 2.4.3. Calculation of CMRO_2_

Concurrently measuring CBF, CBV and BOLD changes allow us to calculate CMRO_2_ according to the Davis model (Davis et al., 1998) using M and β estimated from experiment 2 for each participant:

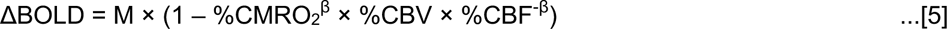

#### 2.4.4. Dynamic signal processing and model fitting for BH hypercapnia (Experiment 2)

Dynamic VASO and BOLD signals were detrended (repeat for each run) to remove low-frequency physiological signal drift (Yan et al., 2009). Detrending was not performed for ASL due to pair-wise subtraction of label and control signals. Time points were identified as outliers if they fell beyond the 15^th^ and 85^th^ percentiles of the BOLD and VASO signals and 25^th^ and 75^th^ percentiles of ASL signals across resting-state and task runs. The corresponding BOLD and VASO signals were capped to the 15^th^ or 85^th^ percentile values, and the ASL signals were capped to the 25^th^ or 75^th^ percentile values, respectively.

To estimate the delay between the onset of BH and vessel dilation, we conducted a 3-min pre-scan using low-resolution BOLD (2.5 mm-iso, TR = 1000 ms). This delay was incorporate into the timing of BH cues during the experiment for each participant (e.g. participants were cued to start BH a few seconds ahead of acquiring BH data). Due to the inherently slow process of BH induced vessel dilation, a mixed physiological state of BH and resting conditions could potentially present throughout our measurements. This overlap might result in elevated resting state signals due to residual hypercapnia effects. To address this issue, a least-square curve fitting algorithm (MATLAB lsqcurvefit function) was employed to fit M, β as well as baseline CBF, VASO and BOLD simultaneously from all time points including all rest and BH scans (5 unknowns and 54 time points).

### 2.5. Segmentation of cortical layers

ASL images were upsampled to a finer grid of 0.25×0.25×0.5 mm^3^ resolution to avoid singularities at the edges in angular voxel space using the AFNI ‘3dresample’ program with linear interpolation (Cox, 1996). Images were upsampled for cortical depth analysis, allowing a large number of voxels to be randomly sampled at different cortical depths so that a continuous laminar profile can be derived. Examples of FT-induced activation maps at the original spatial resolution can be found in Supplemental Figure S2, and double-peak activation patterns are still observable. The borderlines of CSF/GM and GM/WM in M1 hand-knob area were manually drawn on ASL control images. Fifteen cortical layers of M1 were segmented by LAYNII (Huber et al., 2021) using the equi-volume layering approach (Waehnert et al., 2014). Segmentation of cortical layers in V1 was done on reconstructed surface. A boundary-based algorithm was used to co-register structural MP2RAGE UNI images to ASL control images (Saad et al., 2009). MP2RAGE volume was skull removed and segmented into WM, GM and CSF using FreeSurfer (7.1.1). AFNI/SUMA and custom python codes were used to generate the equi-volume surfaces (https://github.com/herrlich10/mripy) between WM and pial surfaces. Eight cortical layers (1 WM, 6 GM and 1 CSF) in V1 were then projected back to volume space for analysis using the AFNI ‘3dSurf2Vol’ program with mapping function ‘max’ (https://afni.nimh.nih.gov/pub/dist/doc/program_help/3dSurf2Vol.html).

### 2.6. Statistical analysis

Paired t-tests were performed to test the significant increase of BOLD, CBF, CBV and CMRO_2_ in the visual stimulus region (4 – 6 degree of eccentricity) compared adjacent eccentricities. Two-peak patterns were observed in FT activation in M1 (CBF and CBV) and fovea inhibition in V1 (CBF and CMRO_2_). To test the significance of the two-peak pattern, we calculated R^2^_diff_ score for each laminar profile (Shao et al., 2021). R^2^_diff_ was defined as the difference between the adjust R^2^ estimated from curve fitting assuming two or single Gaussian distributions. Larger R^2^_diff_ indicates the profile can be better described as two peaks instead of a single peak. Statistical significance of the R^2^_diff_ was estimated by 5000 Boot strapping runs with random noise in each cortical layer estimated by inter-participant variation. P value was calculated as the probability that R^2^_diff_ score of profile can be explained by noise only. Linear regressions were performed to assess the correlations between CBF, CBV, BOLD and CMRO_2_ signals of the visual stimuli experiment. After correcting for multiple comparisons, adjusted P < 0.004 was considered as significant for t-test (12 comparisons), and adjusted P < 0.006 was considered as significant for two-peak detection (8 comparisons).

### 2.7. pRF calculation

First, we sampled the visual field into a 40×40 grid (both the x-axis and y-axis are from - 10 degrees to 10 degrees, with a step of 0.5 degrees). As shown in Supplemental Fig. S4, we obtained the time series of the visual stimulus sweep of each node in the grid. Then we selected a voxel to extract its time series. The time series of this voxel was deconvolved and combined with the sequences of all nodes in the grid to calculate the response of the neurons in this voxel to the visual stimulation at each node. In this way we could get the receptive field map of the neurons in this voxel, then find the center position of the receptive field and calculate its position in the visual field (eccentricity and angle). Finally, we obtained the eccentricity corresponding to this voxel, and calculated the eccentricity map by repeating this step for all voxels.

## 3. Results

### 3.1. Double-peak activation in M1

FT task elicits neuronal activities across both superficial and deep cortical layers: the superficial layers are hypothesized to be engaged by sensory inputs from the somatosensory (S1) and premotor cortices, while the deep layers are hypothesized to be involved in motor outputs (Supplemental Fig. S1). The FT task, commonly utilized in laminar fMRI studies for its efficacy in probing cortical layer-specific activities, served as a key component in our study to validate the sensitivity of our proposed technique. Fig. 3 shows FT-induced signal changes and laminar profiles for CBF, BOLD, VASO and CBV (CBV without BOLD correction was also shown for comparison). Employing this technique, we observed increases in CBF and BOLD signals, alongside decreases in VASO signals along the omega-shaped M1 hand-knob, with double-peak patterns observable upon visual inspection (Fig. 3 Middle). The right panels of Fig. 3 display average laminar profiles from four participants, showing up to 126±22% (Mean±SD) (range: 80±22% to 126±22%) increase in CBF and a 28±2% (range: 19±3% to 28±2%) increase in CBV in the deep layers, and up to 111±31% (range: 62±34% to 111±31%) increase in CBF and a 46±7% (range: 19±4% to 46±7%) increase in CBV in the superficial layers. The observed double-peak activation patterns were statistically significant for both CBF (R^2^_diff_ = 0.79, P < 0.001 for %CBF and R^2^_diff_ = 0.54, P = 0.005 for ΔCBF) and CBV (R^2^_diff_ = 0.75, P < 0.001 for CBV without BOLD correction, R^2^_diff_ = 0.35, P = 0.026 for BOLD corrected CBV (not significant after correcting for multiple comparisons)), while the BOLD response predominantly peaked in superficial layers, attributed to significant venous contamination (R^2^_diff_ = 0.11, P = 0.14). These findings align with our hypothesis and verify the intricate layer-specific neurovascular activations during motor execution tasks.

**Figure 3.**
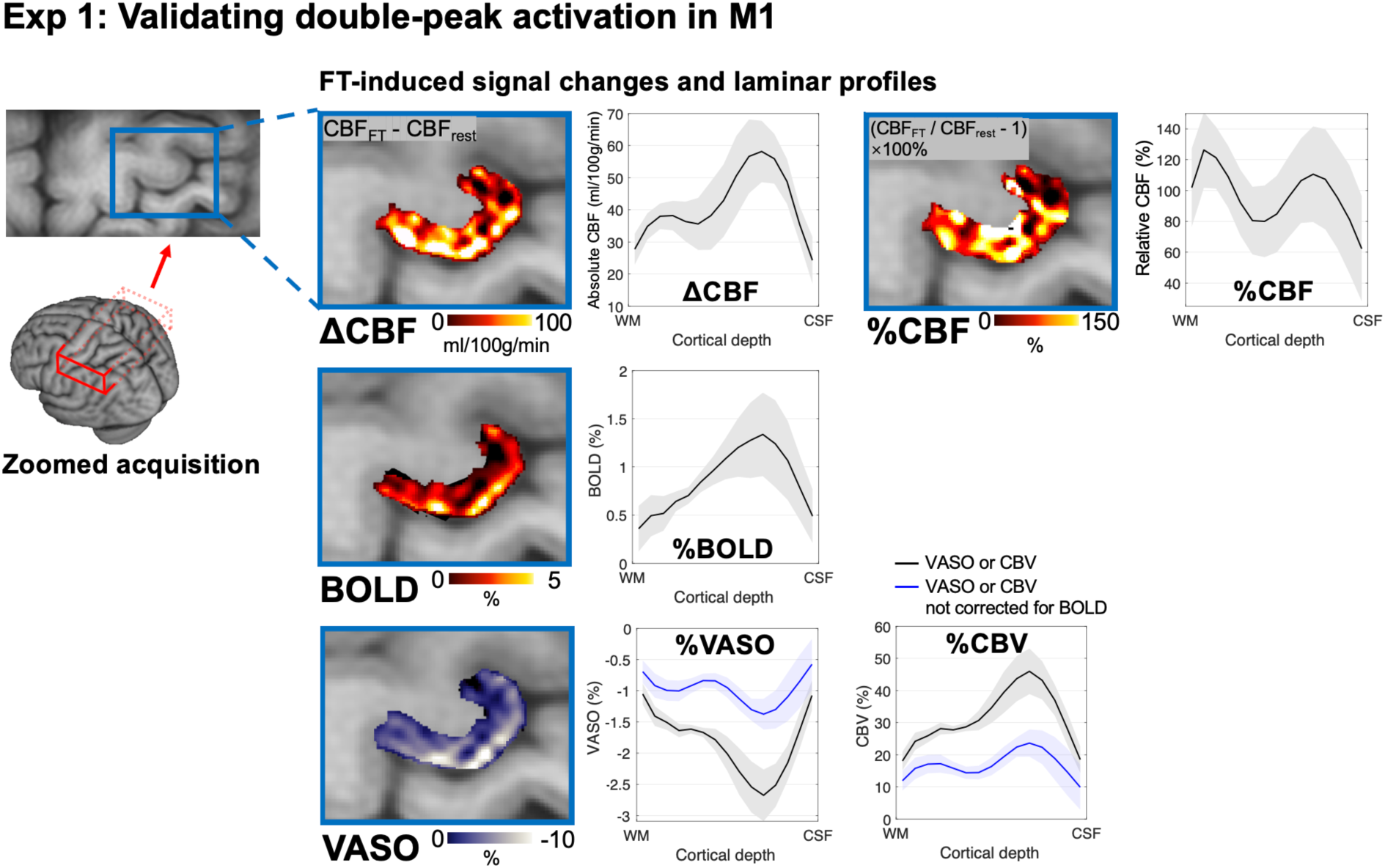
Validation of double-peak activation in the primary motor cortex (Experiment 1). A zoomed field of view (FOV) (80×40×30.8 mm³) was placed to measure the activities in the dominant motor cortex. The experiment consisted of interleaved rest and sequential finger tapping (FT) exercises. FT is expected to induce neuronal activity across both superficial and deep cortical layers, hypothetically engaging proprioceptive and exteroceptive sensory inputs from the somatosensory (S1) and premotor cortex, and motor output from deep layers. The four insets show the FT-induced absolute (Δ) and relative (%) CBF, BOLD, and VASO signal changes and adjacent plots are the laminar profiles of CBF, BOLD, VASO and CBV collated from four participants. These maps were upsampled to a finer grid (0.25×0.25×0.5 mm^3^) for cortical depth analysis, and activation maps at the original spatial resolution can be found in Supplemental Figure S2. The observed ’double-peak’ patterns in CBF/CBV enhancements align with the sensory inputs in superficial layers and motor outputs in deeper layers. Notably, the BOLD signal predominantly peaks within superficial layers, due to the significant contribution from venous contamination. Shaded regions indicate standard error across four participants.

### 3.2. Calibrated fMRI with breath-hold (BH)-induced hypercapnia

Fig. 4 shows neurovascular responses induced by BH hypercapnia of a representative participant. The observed global increases in both BOLD and CBF signals, accompanied by a decrease in VASO signals (indicative of CBV increase), reveal the dynamic nature of blood supply and its regulation in response to elevated CO_2_ levels (Fig. 4 top). The laminar profiles further show an up to 45±16% (Mean±SD) (range 32±16% to 45±16%) increase in CBF peaking in the deep layers and up to a 19±2% (range 14±2% to 19±2%) increase in CBV peaking in the superficial layers (Fig. 4 middle). The observed patterns of BOLD, CBF, and CBV signals indicate a layer-specific vascular response to hypercapnia, implying that the profiles of M and β are also likely to vary across cortical layers. The bottom panel of Fig. 4 shows the laminar profiles of M and β. Our findings provide a detailed laminar profile for both M and β, which enables the calculation of CMRO_2_ across cortical layers in subsequent experiments.

**Figure 4.**
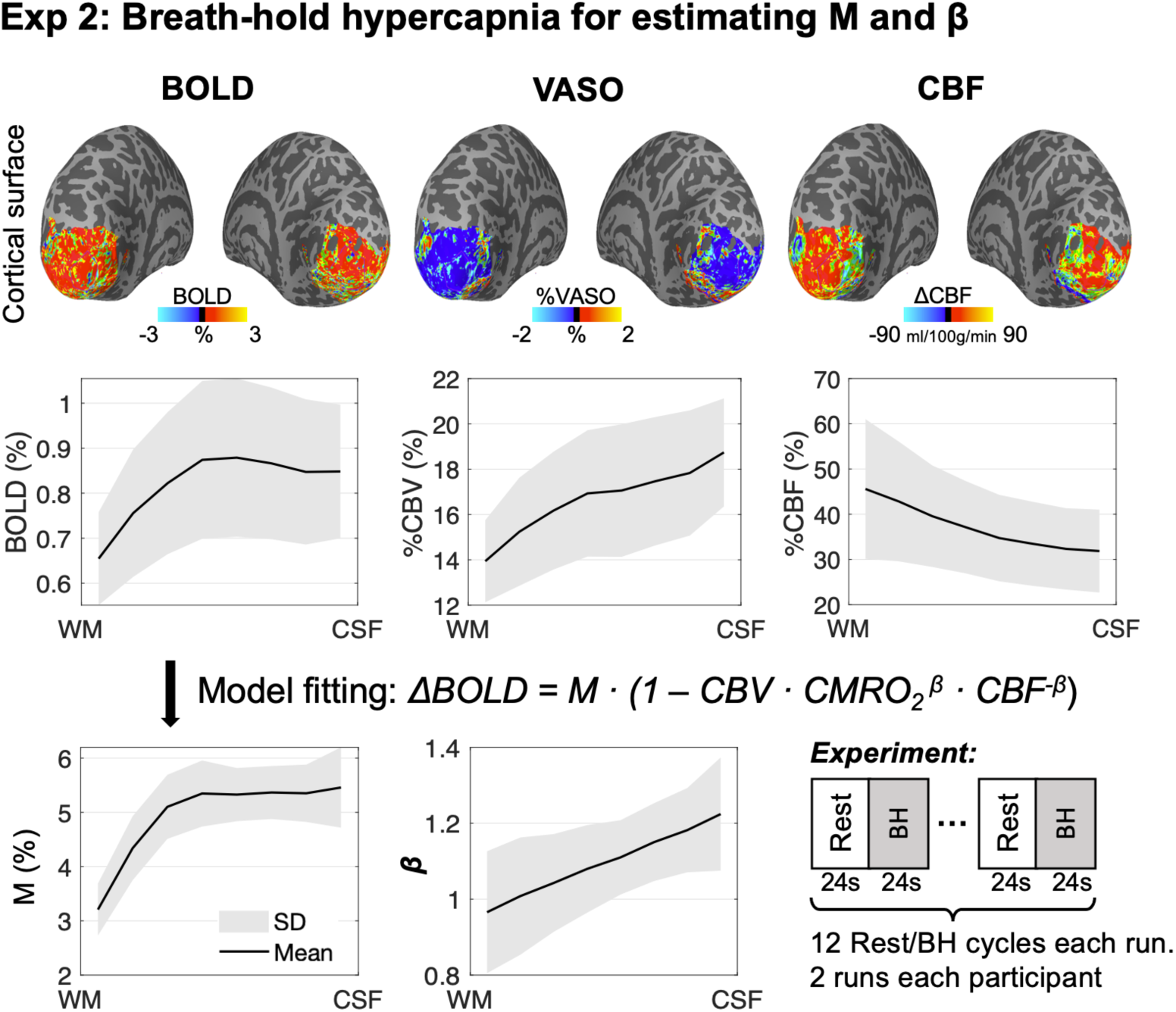
Neurovascular responses to breath-hold-Induced hypercapnia of a representative participant (experiment 2). The first row shows changes in BOLD, VASO and CBF across the cortical surface following BH-induced hypercapnia. A global increase in both BOLD and CBF signals can be observed, as well as decrease in VASO signals (increase in CBV). The second row shows the laminar profiles of BH-induced BOLD, CBV and CBF signal changes. The bottom panel shows laminar profiles of fitted M and β parameters according to the Davis model. The experiment consisted of 12 cycles alternating between 24 sec of rest and 24 sec of BH. Two runs were repeated for each participant to improve measurement reliability. Shaded regions indicate standard error across nine participants.

### 3.3. Distinct neurovascular and metabolic responses across cortical layers and eccentricities in the primary visual cortex

Our visual stimulation experiment utilized a ring-shaped stimulus within the 4-6 degree of eccentricity range (Supplemental Fig. S3) while the participants were asked to perform a color detection task on the central fixation point. We measured layer dependent CBF, CBV, BOLD and CMRO_2_ responses for each eccentricity from 0 to 8 degrees. The definition of ROI is from the pRF experiment (Supplemental Fig. S4). Fig. 5 shows the activation map of a representative participant and the schematic diagram of the ROI definition. Fig. 6A shows the average laminar profile of the changes of CBF, CBV, BOLD and CMRO_2_ responses of nine participants across cortical layers and eccentricity ROIs associated with stimuli, fovea, peripheral and transition regions. For different eccentricity areas, we performed multi-contrast fMRI analysis separately to derive the laminar profiles of CBF, CBV, BOLD and CMRO_2_ for different eccentricity ranges respectively.

**Figure 5.**
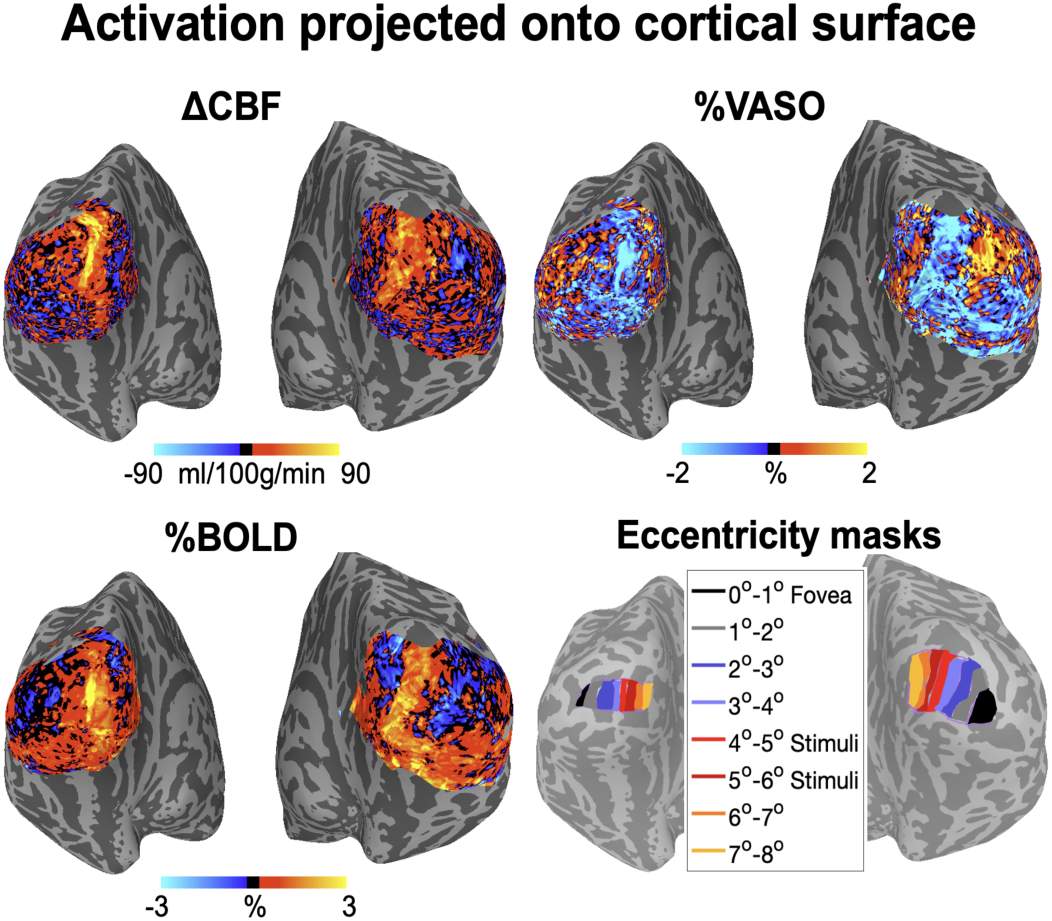
Activation maps of a representative participant with ring-shaped stimulus and eccentricity masks (experiment 4). This figure shows the different neurovascular responses induced by a ring-shaped visual stimulus targeting the 4-6 degree eccentricity range. Within this specified stimulus range, increases in CBF and BOLD signals, and a decrease in VASO signals can be observed. In contrast, mixed or opposite responses are observed in adjacent regions, indicating the localized specificity of the stimulus-induced neuronal and vascular activities.

**Figure 6.**
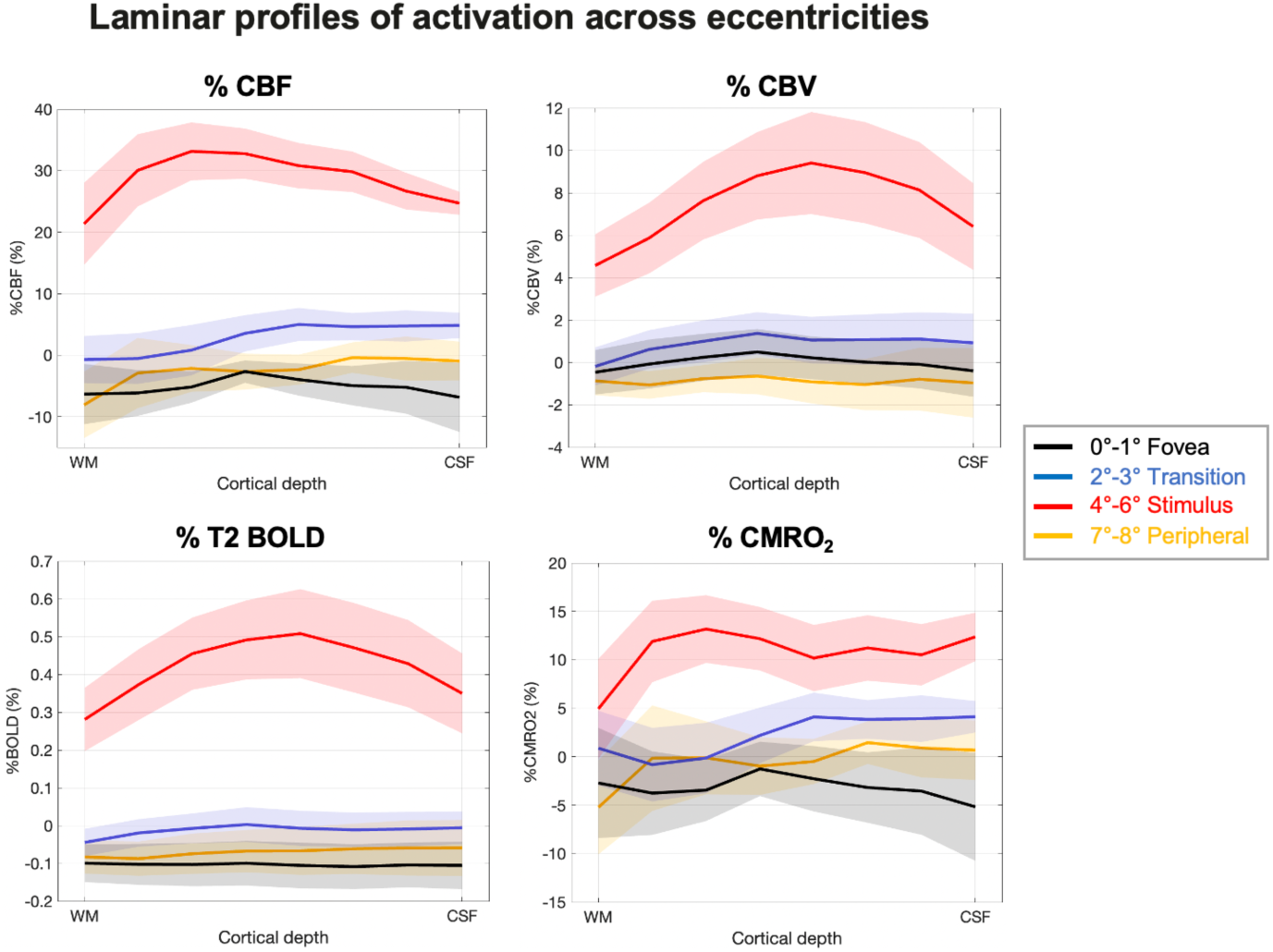
Laminar profiles of CBF, CBV, BOLD and CMRO_2_ signal changes induced by ring-shaped stimulus. Laminar profiles for each degree of eccentricity. CMRO_2_ was calculated from CBF, CBV and BOLD signals incorporating the M and β measured from experiment 2. Shaded areas indicate standard error across nine participants.

#### 3.3.1. Feed-forward visual stimuli (4 - 6 degree of eccentricity)

From the activation maps (Fig. 5), we observed a significant increase in CBF and BOLD signals, and a decrease in VASO within the 4 - 6 degree of eccentricity range. This matches well with the areas that our stimulation activates. We observed that the multi-contrast fMRI responses of the 4 - 6 degree of eccentricity range (Fig. 6A) were significantly greater than the respective responses in other eccentricities (P<0.001 for BOLD, CBF, CBV and CMRO_2_). Although the peak activation from feed-forward stimuli is commonly expected to occur in the middle towards deep layers (Lamme et al., 1998), we observed a slight shift between the profiles of CBF and CBV responses (Fig. 6 top). The peak of the laminar profile in the CBF signal is located in the deeper middle layer, while the peak of the CBV signal is in the more superficial part of the middle layer. Despite the shifted peaks observed in CBV and CBF profiles, CMRO_2_ increase was relatively consistent across cortical layers, with a large peak in the middle to deep layers and two subtle bumps in middle and superficial layers.

#### 3.3.2. Negative BOLD responses in fovea (0 – 1 degree eccentricity)

For the fovea region (0-1 degree eccentricity range), we found a decrease in CBF and BOLD signals, and an increase in VASO signal in the activation map (Fig. 5), consistent with findings of the previous study (de la Rosa et al., 2021). We performed two-peak activation tests on the laminar profile in the fovea region (black lines in Fig. 6A). We found significant two-peak activation patterns in CBF (R^2^_diff_ = 1.09, P < 0.001) and CMRO_2_ (R^2^_diff_ = 0.78, P = 0.003) signals, while CBV (R^2^_diff_ = -0.71, P = 0.41) and BOLD (R^2^_diff_ = -0.38, P = 0.25) decreases were relatively flat across cortical layers. In other words, the decrease in CBF and CMRO2 is greater in superficial and deep layers than in middle layer.

#### 3.3.3. Adjacent region (7 – 8 and 2 – 3 degree eccentricity)

In addition to retinal topographic projection region and fovea, the proposed technique allowed investigation of neuronal activities in peripheral (7-8 degree) and transitional (2-3 degree) regions. Since the fMRI activation patterns of these two areas are relatively similar, we pooled the two together as the adjacent regions to study the activation pattern.

For adjacent regions, it can be clearly seen from the activation map in Fig. 5 that these areas show a decrease in BOLD and CBF signals and an increase in VASO signal (indicative of CBV decrease), which is opposite to the stimulus activation pattern. Furthermore, the laminar profiles of adjacent regions shown in Fig. 6A suggests a potential lateral inhibition mechanism. The CBF and CMRO_2_ signals show a trend of lower signal intensity in the deep layers than in the superficial and middle layers in the eccentricity range of 2-3 degrees and 7-8 degrees.

### 3.4. The CBF signal has the closest trend to the change of the CMRO_2_ signal

Fig. 6 shows that CBF signal changes have the closest trend to the CMRO_2_ signal changes compared to those for CBV and BOLD. We performed correlation analyses between CMRO_2_ and CBF/CBV/BOLD signals across eccentricities and cortical layers (Fig. 7). Compared to CBV (r = 0.156, P = 0.008) and BOLD (r = 0.147, P = 0.013), CBF (r = 0.782, P < 0.001) showed the highest correlation with CMRO_2_ signal.

**Figure 7.**
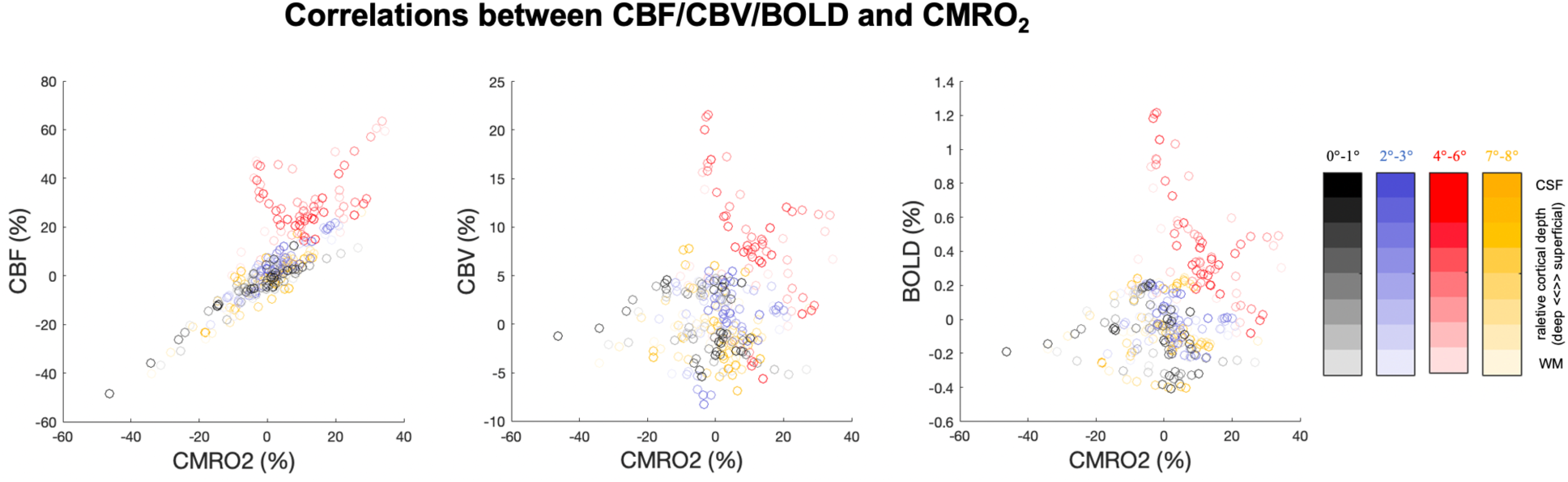
Correlations between CBF/CBV/BOLD and CMRO_2_. Each point represents a signal change at one eccentricity and one cortical depth of one participant. Solid red lines indicate the linear regression results between two measurements. Color bar shows the colors of the signals at different eccentricity and cortical depth.

## 4. Discussion

In this study, we introduced an innovative laminar multi-contrast fMRI technique to study the neuronal mechanisms underlying the positive and negative BOLD responses at 7T. Compared to lower field strengths (e.g. 1.5T or 3T), UHF strengths enable the achievement of mesoscale fMRI at the level of cortical layers and columns (Beckett et al., 2020; Chai et al., 2024; Finn et al., 2019; Huber et al., 2017; Yacoub et al., 2007), primarily due to a superlinear increase in SNR with field strength (Pohmann et al., 2016). UHF ASL benefits from both a significantly higher SNR and a prolonged tracer half-life (Huber et al., 2019; Ivanov et al., 2018; Kashyap et al., 2021; Li et al., 2016; Pfeuffer et al., 2002; Shao et al., 2021). These advantages address the primary limitation of ASL in terms of low SNR, thereby enhancing the spatial resolution and reliability of the CBF measurements. We demonstrated the high specificity in detecting distinct CBF, CBV, and BOLD signal profiles across cortical layers utilizing the FT experiment. The FT task induced double-peak activation patterns in CBF and CBV signals within the M1 hand-knob area, reflecting the underlying neurocircuits involved in receiving and transmitting signals (Huber et al., 2017). However, the exact neuronal mechanisms need further validation. Although animal studies have shown anatomical connections providing evidence for layers involved with cortico-cortical input and cortico-spinal output (Mao et al., 2011; Weiler et al., 2008), Poplawsky et al. measured CBV responses in the rat olfactory bulb with odor and electrical stimulation and found that deep-layer CBV responses can reflect synaptic inputs in the rat olfactory bulb (Poplawsky et al., 2015). Nevertheless, we demonstrated the high specificity of the proposed technique, which is crucial for our study to accurately estimate laminar CMRO_2_ profiles and examine the complex neurovascular and metabolic responses associated with both positive and negative BOLD signals.

There is a limited number of studies utilizing concurrent high resolution CBF, CBV and BOLD measurements to investigate the neuronal activities (Cheng et al., 2017; Krieger et al., 2015; Yang et al., 2004). Moreover, previous calibrated fMRI studies (Blockley et al., 2013; Griffeth & Buxton, 2011; Krieger et al., 2014) generally assumed that changes in CBV and CBF follow Grubb’s power law (CBV ∝ CBF^α^, α = 0.38 (Grubb Jr et al., 1974)). The Grubb’s power law allows CMRO_2_ estimation from BOLD signals using either CBF or CBV data. However, given the complexity of the neurovasculature, the relationship between CBV and CBF becomes complex, particularly when examining neuronal activity in different brain regions (Chen & Pike, 2009). By applying the Grubb’s power law to our concurrent CBF and CBV measurements, we found an average α = 0.36±0.10 across all eccentricities and cortical layers, aligning well with the literature (Grubb Jr et al., 1974). However, notable variations of the Grubb’s constant were observed across cortical layers as well as during specific tasks (Fig. S5 A). Within the stimulation region, α peaks towards superficial layer (∼0.37) while lower α values were found in deep layers (∼0.21), which reflects the differences of the underlying microvasculature (Chen & Pike, 2009). Difference profiles of α values were also observed between the stimulation region and the fovea, which further emphasizes that assuming a constant α value may be inaccurate for assessing the underlying neuronal activities for mesoscopic fMRI studies. In our study, when comparing CMRO_2_ values measured from concurrent CBF, CBV, and BOLD measurements, we observed significant underestimations of CMRO_2_ within the stimulation region by 68.0±40.6% when using BOLD and CBV, or by 27.2±18.5% when using BOLD with only CBF data (Fig. S5 B).

Several approaches were developed to estimate the CMRO_2_ (Lu et al., 2004; Rodgers et al., 2016; Xu et al., 2009), while the calibrated fMRI technique based on the Davis model has been commonly utilized (Davis et al., 1998). This model demonstrates the correlation between CMRO_2_ changes and variations in CBF, CBV and BOLD signals via two parameters, M and β (Davis et al., 1998). M denotes the maximal change in the BOLD signal achievable based on baseline deoxyhemoglobin levels, while β accounts for the impact of localized magnetic susceptibility on R2* differences, emphasizing the vascular distinctions between capillaries and larger vessels, and is expected to change across field strength, TE, and BOLD contrast (T2 versus T2*). Commonly, M and β can be estimated under the assumption that the CMRO_2_ remains constant during hypercapnia. However, the slow dilation of vessels induced by breath-holding (BH) may lead to a mixed physiological state between BH and resting-state measurements. We adjusted acquisition timing based on BH-signal delay measured from a pre-scan, however, resting state CBF, CBV and BOLD signals can still be over-estimated due to residual hypercapnia. To mitigate this delay and potential fluctuations of BH hypercapnia levels, we considered all scans as BH condition (i.e. resting-state signal is zero to low-level BH) and simultaneously estimated M, β, as well as baseline CBF, CBV and BOLD values from all measurements. Supplemental Fig. S6 shows an example of dynamic CBF, CBV, and BOLD signals from experiment 2. Despite signal fluctuations across the measurements, our method can effectively predict BOLD signals using measured CBF and CBV, along with estimated M and β values and RSME between measured and predicted BOLD signals was 0.0050±0.0019 across all participants and cortical layers. BH-induced CMRO_2_ changes were averaged to be 0.3% (ranging from -0.96% to 2.39%) across eight cortical layers (Eq. [5]), which indicates minimal CMRO_2_ variation due to BH and further validates the M and β measurements.

In this study, the observed patterns of BOLD, CBF, and CBV signals responses induced by BH hypercapnia indicate a layer-specific vascular response to hypercapnia, allowing us to measure detailed laminar profiles for M and β (Fig. 4). This layer-specific vascular response may reflect the complex architecture of cortical vasculature probed by BOLD, VASO and ASL. Since GRASE acquisition was utilized to generate T2-weighted BOLD contrast, we observed peak of BOLD increase in the middle layer. While gradient echo (GE) BOLD signal scales with vessel diameter and is predominantly sensitive to larger draining vein near cortical pial surface (Siero et al., 2013), T2 BOLD signal is generally believed to reflect the micro-vasculature at UHF (Markuerkiaga et al., 2016; Uludağ et al., 2009), so that observed BOLD signal is less biased by the venous contamination in the superficial layers. CBV changes reflect hypercapnia-induced vasodilation in both macro- and micro-vasculature, where CBV sampled from macro-vasculature (arterioles) are hypothesized to increase from deep to superficial layer but not for the micro-vasculature compartment (capillaries) (Schellekens et al., 2023). ASL signal primarily originates from capillary vessels, and peak of CBF increase in the deep layer is consistent with the higher microvascular density in middle to deep cortical layers and relatively lower resting state CBF in deep layers (Shao et al., 2021). With the concurrent measurement of BOLD, CBV and CBF, we were able to derive laminar profiles for M and β. For the laminar profile of M, we observed a plateau from middle to superficial layers with relatively lower amplitudes (∼5.5%) compared to EPI BOLD signals (∼14%) (Guidi et al., 2016; Schellekens et al., 2023). To the best of our knowledge, no study has reported detailed laminar profiles of β at 7T, while β is usually assumed to be 1 for UHF studies when the venous signal becomes negligible in extravascular BOLD signals (Griffeth & Buxton, 2011; Kida et al., 2000; Krieger et al., 2014). However, β is associated with localized magnetic susceptibility on R2* differences and is likely to change when different TE or BOLD contrasts are used (T2 versus T2*). We found β gradually increases from deep (0.97) to superficial layer (1.22) with an average value of 1.10±0.09 across layers. Compared to GE-BOLD, GRASE BOLD is more sensitive to the underlying microvasculature (Scheffler et al., 2021), which is likely to change across cortical layers. Thus, measuring laminar profiles of M and β provides insights into the underlying neurovascular coupling mechanisms and is crucial for accurate CMRO_2_ estimations in laminar fMRI studies.

Based on observed CBF, CBV, BOLD and CMRO_2_ responses induced by partial visual field activations stimulus, Fig. 8 shows our hypothesized neural circuits for the stimulated area, peripheral and transition area, and the fovea of V1, along with associated layer-dependent response patterns.

**Figure 8.**
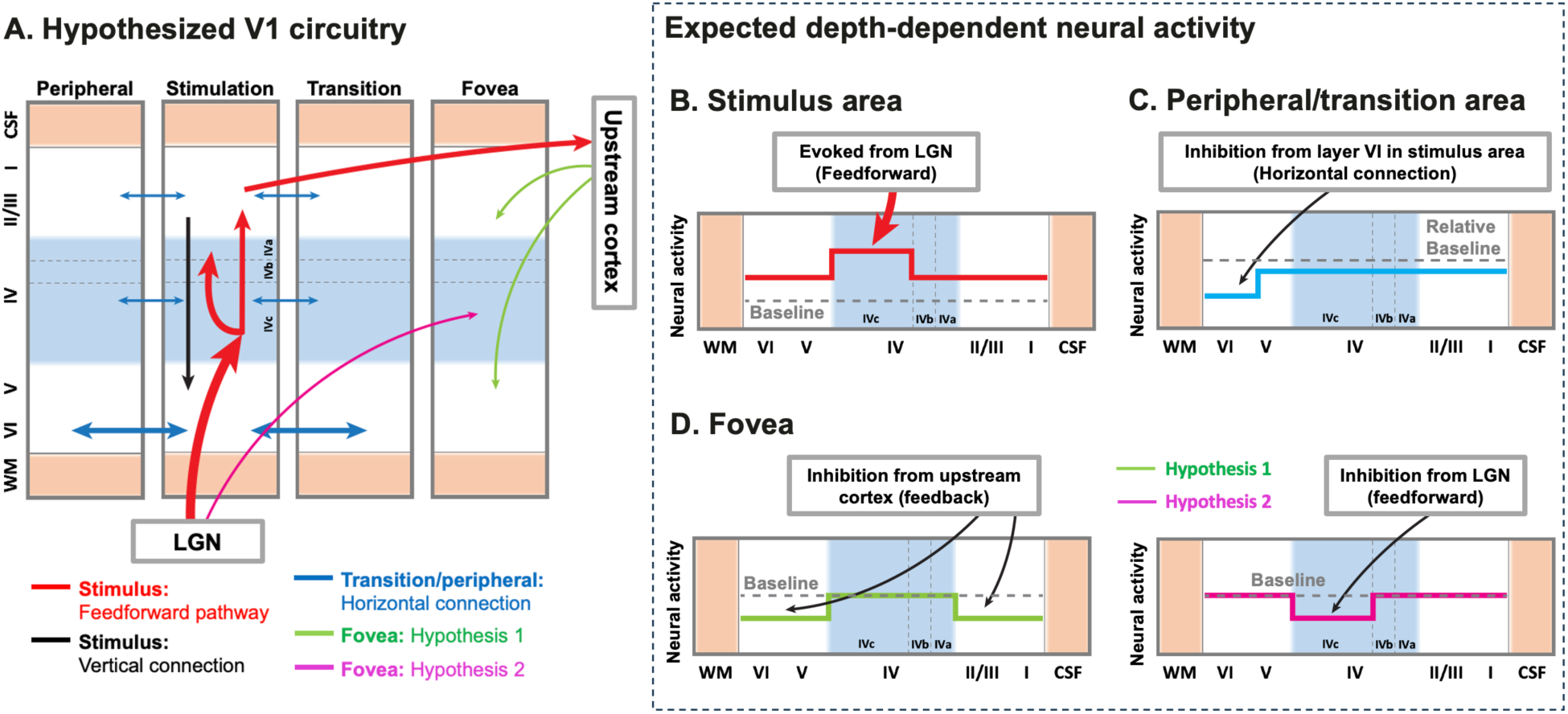
Hypothesized models of the laminar circuitry in V1. (A) The model of layer-dependent circuitry based on the literature. The red arrows show the hypothesized feedforward pathway through which visual information is transmitted from LGN to V1 and then downward. The black arrow represents the vertical connection between V1 layers. The blue arrows represent the horizontal connections between different regions in V1. Previous research has shown that the deeper layers (layer VI) have stronger and more extensive horizontal connections (Liu et al., 2014; Srivastava et al., 2022). The green and pink arrows respectively represent our two hypotheses for the negative signals in the fovea. (B) Expected depth-dependent activity in stimulated area with a stronger activation in the middle layers when the stimulus is presented (red trace). (C) Expected depth-dependent activity in peripheral and transition areas. These areas are mainly inhibited by horizontal connections from layer VI of the stimulated area during stimulus presentation (blue trace). (D) Expected depth-dependent activity in the fovea. Inhibition in the fovea is hypothesized to originate from the feedback pathway of upstream cortex (Hypothesis 1, green trace) or through the feedforward pathway of LGN (Hypothesis 2, pink trace). Gray dashed traces indicate baseline (zero activity) in (B, D) and relative baseline comparing to normal activity in (C).

For the visual stimulation region in V1, activation map and laminar profile across participants show the strongest response and a pattern where middle layer has a stronger response than superficial and deep layers. It is known that this neural activity mainly arises from the feedforward pathway of the LGN. Sublamina 4Cα receives mostly magnocellular input from the LGN, while layer 4Cβ receives input from parvocellular pathways. After processing, the signal is transmitted to layers 4A/B and 2/3. Layer 2/3 then sends afferents to layer 5/6 and the higher-level visual cortex (Fig. 8A., Accordingly, we predicted that this neural activation should show a pattern of greater response in the middle towards deep layer (4) with moderate increase in superficial (4A/B and 2/3) and deep layers (2/3) (Fig. 8B). While our results matched this hypothesis very well, we observed a slight shift between the profiles of CBF and CBV responses (Fig. 6 top). Specifically, for CBV measured by VASO, the signal changes reflect contributions from all vasodilation vessels associated with neuronal activity, including pial vessels, penetrating arterioles and capillaries (Huber et al., 2019). We observed that CBV increase peaks towards the mid-to-superficial layers, suggesting that pial arteries and penetrating arterioles in the mid-to-superficial cortex may exhibit greater vasodilatory responses compared to those in deeper layers (Duvernoy et al., 1981). The signal of labeled blood in ASL is considered to primarily originate from capillaries. We employed pCASL with higher SNR than pulsed ASL (Huber et al., 2019; Kashyap et al., 2021). We have measured the arterial transit time (ATT) using a multi-PLD approach with the proposed labeling scheme and zoomed GRASE acquisition (Shao et al., 2021), which was found to range from 0.9 to 1 sec from superficial to deep layers in V1, and applied sufficiently long PLD to minimize pial arterial artifacts (Alsop & Detre, 1996; Alsop et al., 2015), thereby offering a “clean” depiction of CBF changes within the capillaries that are closely related to neuronal activation. The laminar profile of CBF activation indicates greater increase within the mid-to-deep layers compared to baseline, which is consistent with previous laminar perfusion fMRI studies (Kashyap et al., 2021; Shao et al., 2021) and findings from animal research on microvascular density (Jin & Kim, 2008). However, CMRO_2_ increase was relatively consistent across cortical layers, with one large peak in middle to deep layers and two subtle bumps in middle and superficial layers, while a similar pattern was observed in the visual cortex of monkeys in a recent calibrated fMRI study (Bohraus et al., 2023). The laminar profile of CMRO_2_ suggests the feed-forward visual stimuli involve the primary visual signal from the LGN predominantly targeting layer IV, with slight extensions into layers V and VI (Clay Reid & Alonso, 1995) and the potential projections from layer IV to superficial layers (Callaway, 2004) (Fig. 8A, B).

For the fovea region, we found that the decrease in CBF and CMRO_2_ is significantly greater in superficial and deep layers than in middle layer, while BOLD signal shows no difference between layers. The reason for the lack of such results for the BOLD signal may be due to its lower layer specificity (Huber et al., 2019). Previous studies reported negative BOLD responses in the V1 fovea region when visual stimuli were presented in the peripheral visual field (de la Rosa et al., 2021). In this study, participants were asked to perform a color judgment task on the fixation point. Previous research has indicated that when an irrelevant distractor is presented with a target stimulus, the capture of attention by the distractor impairs processing of the target as reflected by slower reaction times to the target (Schreij et al., 2008; Theeuwes, 1992). Moreover, a study by Theeuwes et al (1998) has shown that the attention can also be captured by a new-appearing irrelevant distractor (Theeuwes et al., 1998). There is also evidence from fMRI and electrophysiological experiments that interfering stimuli (destractors) can automatically capture attention and thus affect the processing of task stimuli (Hickey et al., 2006; Spinks et al., 2004). However, the exact neural pathways underlying this interference remain unknown. Interfering stimuli will engage participants’ bottom-up attention processes and thus interfere with participants’ performance on task stimuli. A saliency map is the bottom-up contribution to the deployment of exogenous attention. The mainstream view is that a salience map should integrate various visual information and be formed in the high-level cortex (Itti & Koch, 2001). In contrast, Li proposed that V1 creates a salience map (Li, 2002). Previous studies have indeed found that V1 (Zhang et al., 2012) and superior colliculus (SC) (White et al., 2017) can spontaneously generate a salience map. In view of these two perspectives, we propose two hypotheses (Fig. 8C). One possibility is that attention decreases through feedback connections from upstream cortex, cortico-cortical feedback (Felleman & Van Essen, 1991) (Hypothesis 1 in Fig. 8) targeting the superficial and deeper layers. Another possibility is a SC->TRN->LGN->V1 feedforward pathway (Kolmac & Mitrofanis, 1998; McAlonan et al., 2008) (Hypothesis 2 in Fig. 8) which involves decreased neuronal activity in the middle layer. With the estimation of the laminar profile of CMRO_2_, which is considered a surrogate of neuronal activity, we find that the negative BOLD response in the fovea of V1 is more likely to originate from neuronal inhibition through a feedback pathway from higher visual areas. Specifically, the presented interference stimulus potentially integrates a salience map in high-level brain areas, and then inhibits the original attention response of the primary visual cortex task area through the feedback pathway.

For the adjacent regions, the distance between these regions and the stimulus activation region cannot be ignored, we therefore expect that they will be minimally affected by lateral inhibition when activated by visual stimulation. Previous study used injections of the CVS-11 strain of rabies virus to examine disynaptic long-range horizontal connections within macaque monkey V1 and demonstrated that long-range horizontal connections mainly occur on layer 6 and are responsible for integration across a large region of the visual field (Liu et al., 2014). A recent study found neurons corresponding to this integration in layer 6 of V1 mainly function as GABA inhibitory neurons (Srivastava et al., 2022). Therefore, we predicted that these two areas would exhibit suppressed activation in deep layer when activated by visual stimulation (Fig. 8D). Activation map (Fig. 5) and laminar profile (Fig. 6) clearly show that deep layer CBF and CMRO_2_ signals are more inhibited than superficial layer. Meanwhile, the BOLD and CBV signals do not show this obvious trend. The high resolution of the proposed technique allowed investigation of hemodynamic and metabolic activities within the transitional and peripheral areas of positive and negative BOLD activations. To the best of our knowledge, such investigation hasn’t appeared in literature.

In this study, we found that the CBF (r = 0.782) signal has the highest correlation with the CMRO_2_ signal compared to CBV (r = 0.156) and BOLD (r = 0.147) signals (Fig. 7). Synchronous increase or decrease in CBF and CMRO_2_ were found in 81.6% of all ROIs across eccentricities and cortical layers, while only 53.1% and 47.9% of ROIs had the same direction of CBV and BOLD changes along with CMRO_2_. These results suggest CBF could be the best proxy to CMRO_2_. But actually, the spread across points (different eccentricity, depth, and participants) might contain true spatial patterns as well as inter-participants variability (Lu et al., 2008). Therefore, we used a linear mixed model (LMM) to evaluate the impact of this random effect and the extent to which CBF, CBV, and BOLD signals respectively affect the CMRO_2_ signal.

We performed a z-score for CBF, CBV, BOLD, and CMRO_2_ data points. We then built a LMM to analyze which parameter had the greatest impact on CMRO_2_ and to determine whether differences in spatial location would affect the estimates. The established model is as follows: CMRO_2_ ∼ 1 + CBF + CBV + BOLD + (1|spatial location). The results show that the model has a high fit quality (AIC: -198.08; BIC: -176.1; Loglikelihood: 105.04; Deviance: -210.08). The results of fixed effects coefficients show that CBF, CBV and BOLD all contribute significantly to the fit of CMRO_2_ (intercept/CBF/CBV/BOLD: estimate = 0/1.4325/-0.4402/-0.5159, tStat = 0/96.93/-31.90/-35.78, intercept P = 1 and others P < 0.001). And the contribution of CBF is significantly greater than CBV and BOLD. The random intercept effect of the spatial location group was 0.098813, indicating that there was variability in the effects of different spatial locations on CMRO_2_. However, we noticed that the influence coefficient of CBV and BOLD on CMRO_2_ is negative. We simulated a set of data based on our data distribution range and a similar phenomena can be observed adding noise to the simulated data (Supplemental Fig. S7). In conclusion, combined with the results of this analysis, we can infer that although spatial localizer can indeed influence CMRO_2_ estimate, CBF can still be the best proxy to the underlying neuronal metabolic activity (Gsell et al., 2000).

There are several limitations in this study. The TR was relatively long due to the use of 1280 msec pCASL labeling with a 1160 msec PLD, limiting the capability to capture dynamic responses of CBF, CBV, and BOLD signals (Kashyap et al., 2021; Kim & Kim, 2010). Despite this, pCASL offers advantages including enhanced SNR and less contamination from pial arterial signals compared to pulsed ASL (Alsop et al., 2015), providing higher specificity in laminar CBF profiles (Shao et al., 2021). Future studies might benefit from exploring a dithered acquisition scheme to improve temporal resolution (Kim et al., 2020). To address the inherently slow process of BH-induced vessel dilation and mixed physiological state of BH and resting conditions throughout the measurements in experiment 2. We proposed to fit baseline CBF, VASO, and BOLD simultaneously while fitting M and β values from all time points. Increasing the duration of the resting-state blocks could minimize the residual hypercapnia effect, but this will require accelerated acquisition to improve temporal resolution to capture physiological states more precisely and maintain sufficient BH blocks. Additionally, we employed a single-shot GRASE acquisition method to improve acquisition efficiency, however, relatively thick slices (2.2 mm) were used to provide sufficient spatial coverage. For visual experiments, imaging planes were placed nearly parallel to the calcarine sulcus to preserve high spatial resolution (1 mm) across cortical layers in axial planes, which allows to detect ∼2-3 independent voxels across V1 cortical layers (Fischl & Dale, 2000). However, higher spatial resolution along the slice direction would be desirable to improve laminar measurements and minimize potential signal blurring due to the shift of cortical layers across slices. The specificity of the multi-contrast fMRI can be improved with higher spatial resolution utilizing accelerated acquisition and constrained reconstructions (Chang et al., 2017; Park et al., 2021; Spann et al., 2020; Zhao et al., 2023). It is worth noting that the Davis model is based on a mass-balance principle, meaning that changes in CBV, CBF, and BOLD signals should originate from the vasculature within the voxel being acquired. For mesoscale fMRI, it is possible that oxygenation supply (i.e., through CBF) occurs at different layers than the draining (i.e., BOLD response), potentially violating the assumptions of the Davis model. In this study, we utilized pCASL with optimized PLD and GRASE BOLD with T2 contrast, so that the CBF and BOLD responses are sensitive to more localized laminar microvascular responses and less biased by penetrating or pial macro-vessels. Therefore, applying the Davis model can be meaningful at a higher spatial resolution, as the mass-balance principle hold valid. Regarding CBV measurement, VASO was acquired with GRASE readout with a relatively short TE (17.2 ms), and the acquired CBV is likely to include both arterial (CBV_a_) and venous (CBV_v_) compartments (Huber, Goense, et al., 2014). However, CBV_v_ changes, which contribute most to the BOLD signal, should be included in the Davis model. Incorporating total CBV in the Davis model is likely to underestimate CMRO_2_ due to likely larger CBV responses in arteries (Kim & Kim, 2011). For mesoscale fMRI, the difference between changes in total and venous CBV is expected to be smaller than at coarser scale that includes a mixture of large and small vessels. Guidi et al. proposed estimating CBV_v_ by scaling total CBV with a power of approximately 0.53 (CBV_v_ = CBV_total_^0.53^) (Guidi et al., 2016). This adjustment should be incorporated in future studies to improve the accuracy of CMRO_2_ estimation. However, the scaling factor needs to be verified for the current acquisition parameters as well as with different stimuli (e.g. breath hold versus visual stimulus, as subtle changes in the scaling factor can significantly affect the adjusted CBV_v_, β, and corresponding CMRO_2_ (Supplemental Figure S8). Besides neurovascular responses, BH may decrease arterial oxygen saturation and may affect T2 and corresponding VASO, ASL and BOLD signals. An animal study reported that stimulus-evoked absolute fMRI measurements were relatively invariant across moderate physiological perturbations (i.e. 5% CO_2_), but substantially affected by more extreme physiological perturbations (i.e. 10% CO_2_) (Sicard & Duong, 2005). The potential discrepancies between BH calibration and stimulus evoked CMRO_2_ changes need to be further validated in future studies, especially for mesoscale fMRI where arterial contributions vary across cortical layers. Additionally, we utilized a constant baseline CBV of 0.055 ml/ml across cortical layers (Lin et al., 2008; Lu et al., 2005), but its laminar-dependent variations can directly impact CBV responses and corresponding CMRO_2_ calculations. According to animal and simulation studies (Akbari et al., 2023; Zhao et al., 2006), using a constant total CBV across layers can lead to ∼25% underestimation of CBV change in deep layers and ∼25% overestimation of CBV change in superficial layers. This, in turn, can cause up to a ∼25.7% overestimation of CMRO_2_ in deep layers (β = 0.97) and a ∼20.0% underestimation of CMRO_2_ in superficial layers (β = 1.22). This inaccuracy in CMRO2 can be further amplified when adjusting for CBV_v_ as well as β values. Thus, accurate estimation of baseline CBV is crucial for accurate and quantitative CMRO2 calculation in future studies. Another limitation is the lack of direct validation of our observations, such as the behaviors within the fovea region through methods such as positron emission tomography or electrophysiological recordings (Hickey et al., 2006; McGregor et al., 2020; Poplawsky et al., 2015; Spinks et al., 2004). The distinctive patterns observed in CBF, CBV, BOLD, and CMRO_2_ across cortical layers and eccentricities need to be validated in the future studies to associate with underlying neuronal activities, especially for relatively weak and negative responses in the fovea.

In conclusion, we presented an innovative multi-contrast laminar functional MRI technique that offers comprehensive and quantitative imaging of neurovascular and metabolic responses across cortical layers at 7 Tesla. We demonstrated that the proposed sequence provides quantitative CBF, CBV, BOLD and CMRO_2_ measurements with high resolution and specificity to delineate distinct neurovascular and metabolic responses across cortical layers and eccentricities in the primary visual cortex. This novel technique allows the differentiation of neuronal excitation and inhibition underlying positive and negative BOLD responses, illustrating the associated neural circuit activities across cortical layers.

## Data Availability

The data presented in this manuscript can be requested by contacting the corresponding author. The pulse sequence will be available after we establish Material Transfer Agreement (MTA) between user's institute and University of Southern California. MATLAB code for two-peak pattern significance test can be downloaded from our lab website (http://loft-lab.org/index-5.html).

## 5. Acknowledgement

This work was supported by National Institute of Health (NIH) grant UF1-NS100614, S10-OD025312, R01-NS114382, R01-EB032169, RF1AG084072, R01-EB028297 and R01-NS121040.

## 7. Author contributions

Xingfeng Shao: Conceptualization, Investigation, Conducting experiments, Formal analysis, Writing – original draft. Fanhua Guo: Conducting experiments, Formal analysis, Writing – original draft. JungHwan Kim: Resources, Writing – original draft. David Ress: Resources, Writing – original draft. Chenyang Zhao: Resources, Conducting experiments, Writing – original draft. Qinyang Shou: Resources, Conducting experiments, Writing – original draft. Kay Jann: Resources, Writing – original draft. Danny J.J. Wang: Conceptualization, Supervision, Writing – original draft.

## 9. Supplemental information

**Supplemental Figure S1:**
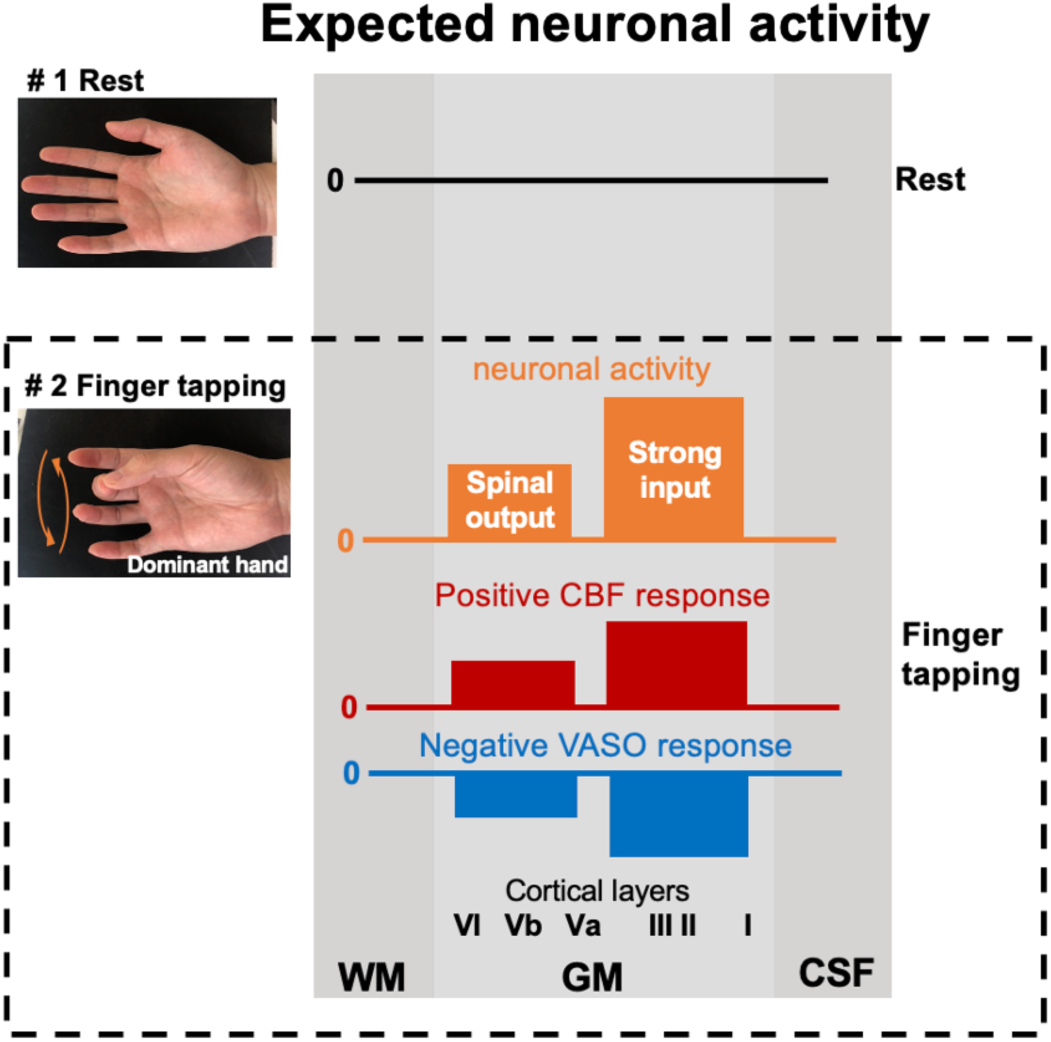
Expected neuronal activation and corresponding CBF and VASO responses to FT. FT is expected to induce neuronal activation across both superficial and deep cortical layers, hypothetically engaging proprioceptive and exteroceptive sensory inputs from the somatosensory (S1) and premotor cortex, and motor output from deep layers. This neuronal activation is expected to associate with double-peak increase in CBF and decrease in VASO (increase in CBV), with stronger responses near the superficial layers.

**Supplemental Figure S2.**
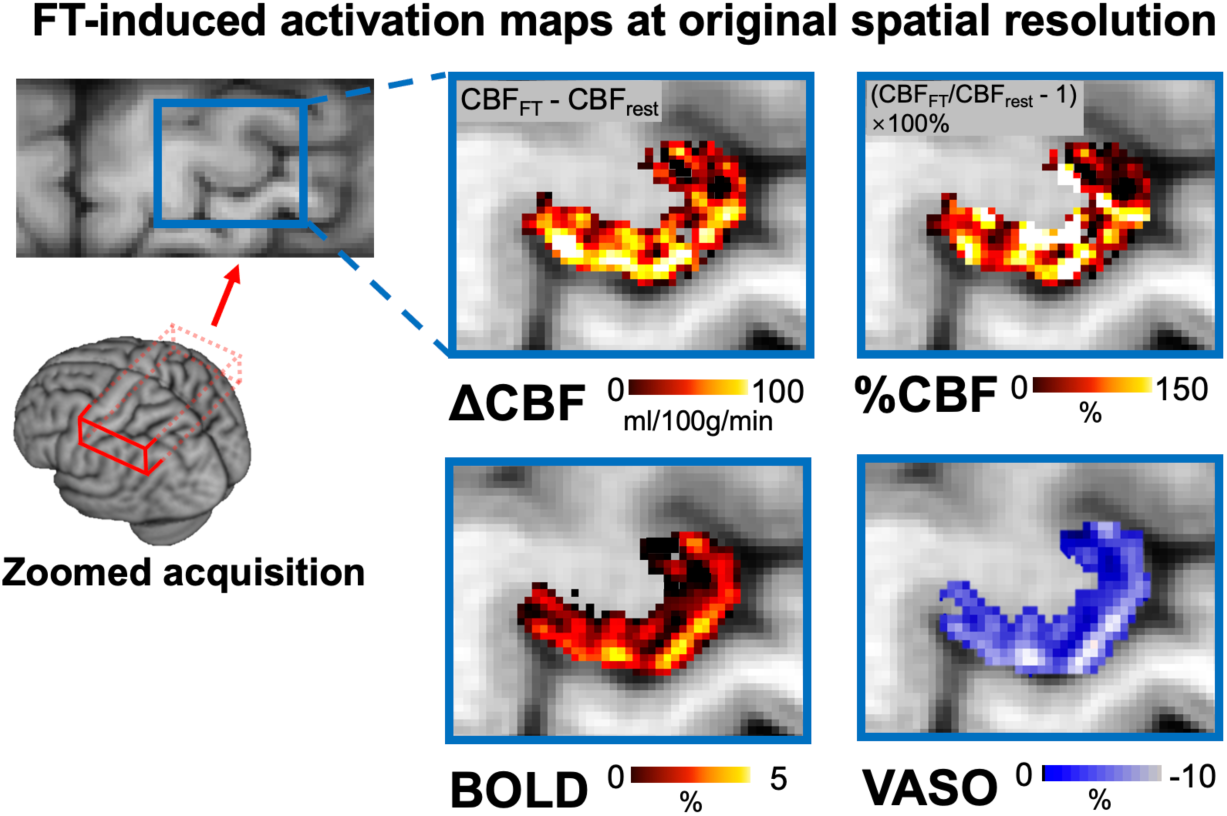
FT induced activation maps at original spatial resolution. These activation maps were shown at the original spatial resolution (1 mm in-plane) before being upsampled to a finer grid for cortical depth analysis. Double-peak activation patterns are still observable at this spatial resolution.

**Supplemental Figure S3.**
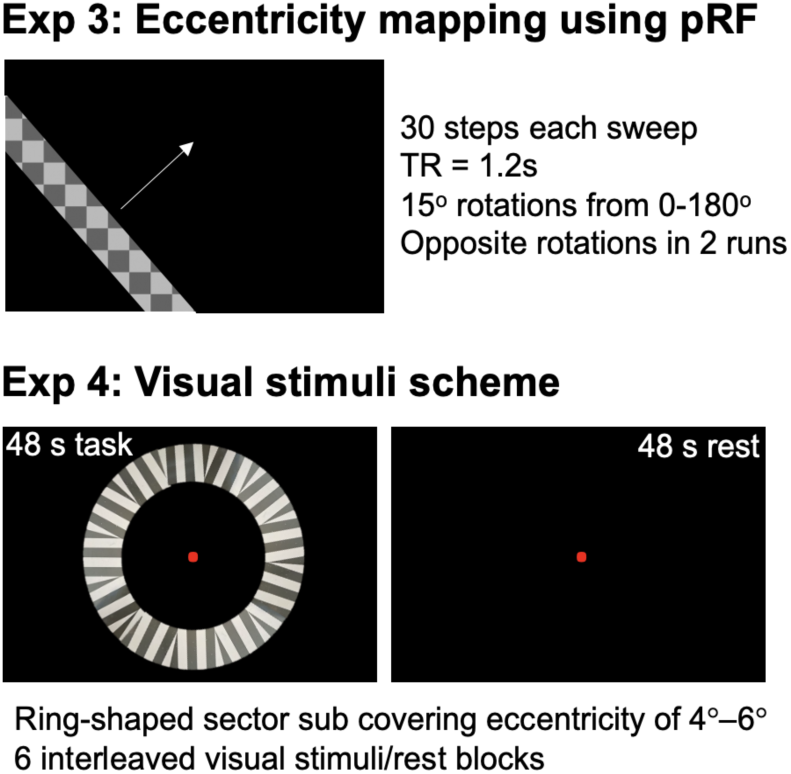
Stimulus schemes for visual stimuli experiments. Top panel shows pRF stimulus pattern for eccentricity mapping (experiment 3): flickering checkerboard pattern that sweeps (30 steps, TR=1.2s) across the screen, sequentially stimulating different visual field regions with a 15-degree rotation after each sweep. The bottom panel shows visual stimulus consisting of a mean-gray background and a ring-shaped sector covering an eccentricity of 4°–6°. Each subsector contained a high-contrast (100%) radial grating (1 cpd) that reversed contrast at 4Hz. One acquisition block (48 sec) acquired two ASL and VASO images and four BOLD images.

**Supplemental Figure S4.**
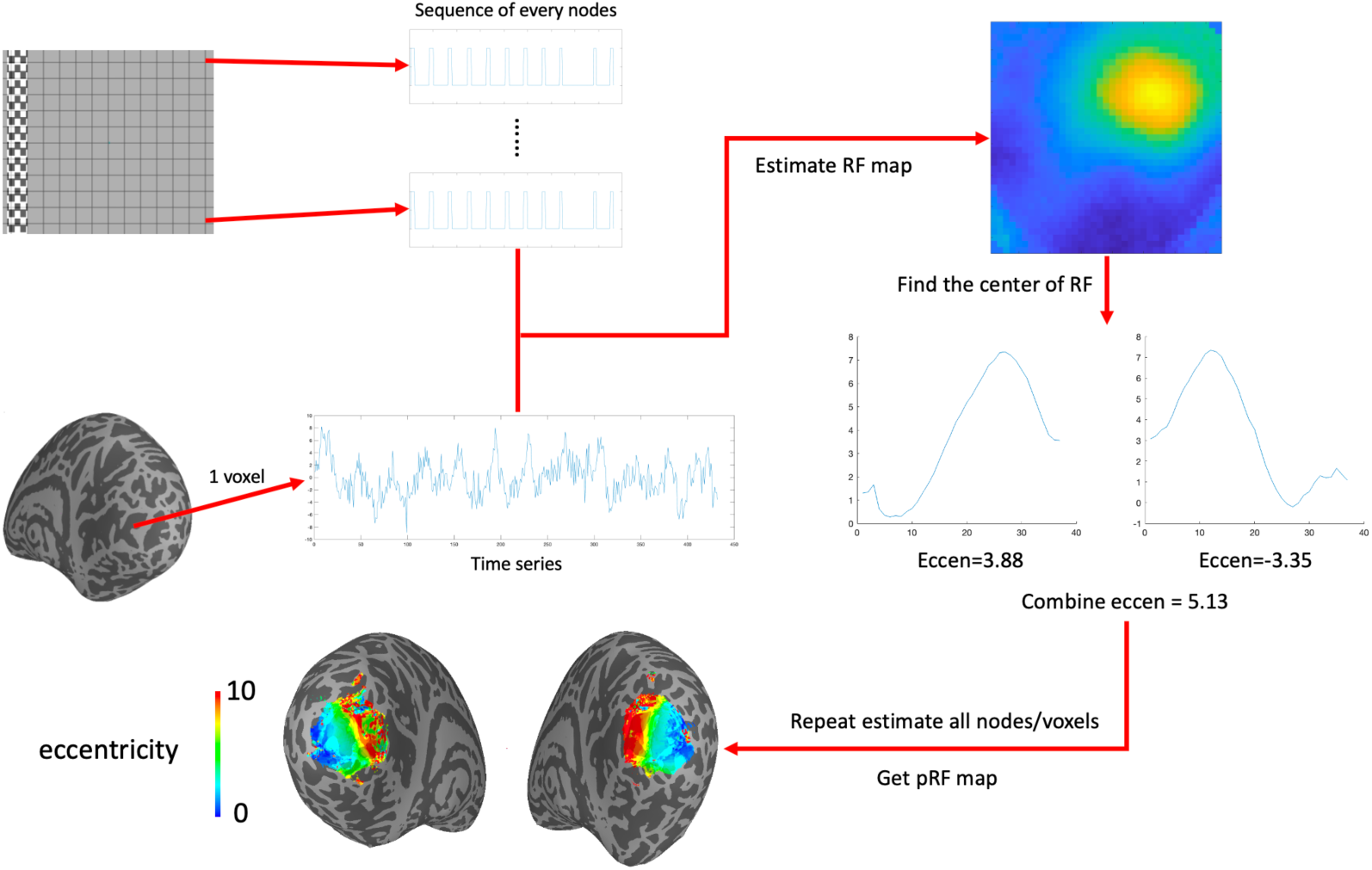
pRF calculation process. First, we sample the visual field into a 40×40 (both the x-axis and y-axis are from -10 degrees to 10 degrees, with a step of 0.5 degrees). As shown in S3, we can obtain the time series of the visual stimulus sweep of each node in the grid. Then we select a voxel to extract its time series. The time series of this voxel is deconvolved and combined with the sequences of all nodes in the grid to calculate the response of the neurons in this voxel to the visual stimulation at each node. In this way we can get the receptive field map of the neurons in this voxel. Then find the center position of the receptive field and calculate its position in the visual field (eccentricity and angle). Then, we can get the centrifugation corresponding to this voxel. We can calculate the eccentricity map by repeating this step for all voxels.

**Supplemental Figure S5.**
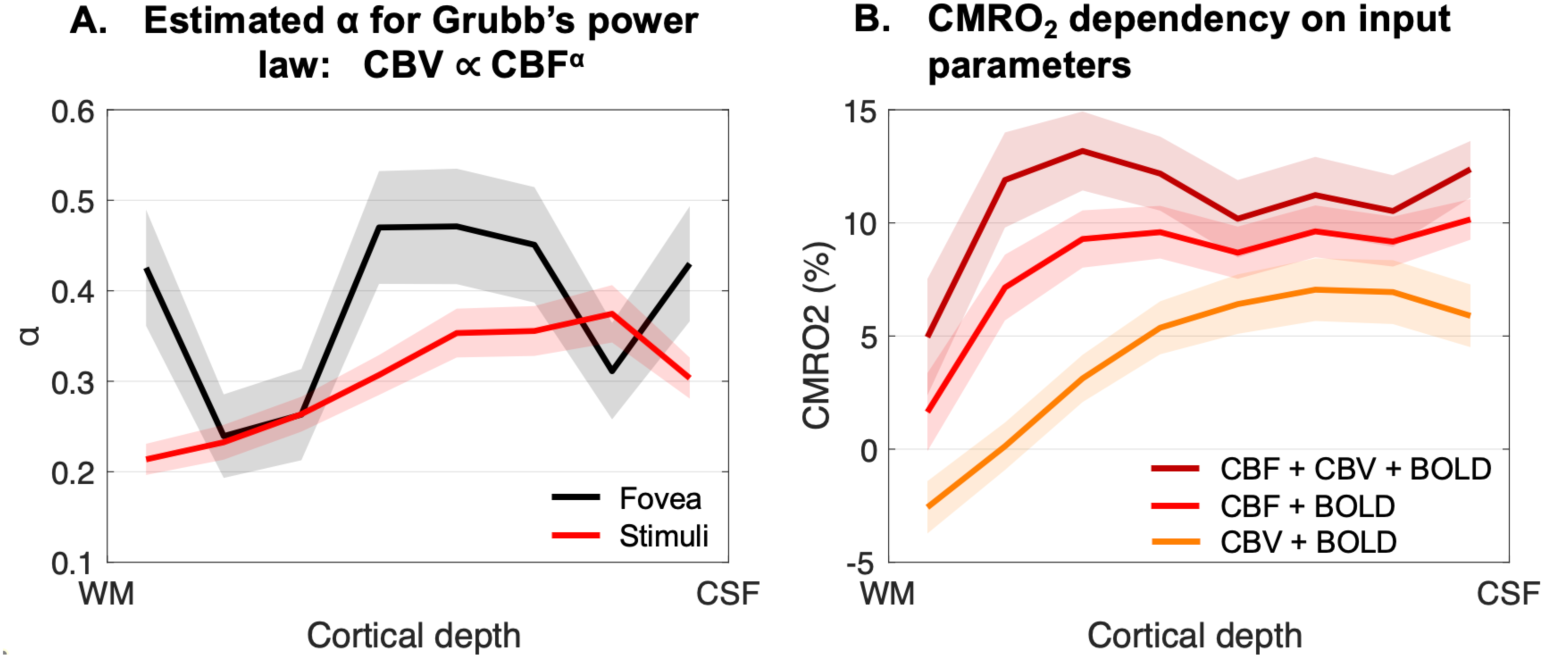
A. Estimated α values in the stimuli and fovea regions using CBF and CBV measurements. B. Comparisons between CMRO2 measured from BOLD and simultaneously measured CBF and CBV (reference, dark red trace), or with only CBF (light red trace) or CBV (orange trace). When only CBF or CBV was used for CMRO2 calculation, the Grubb’s power law was utilized to generate the corresponding CBV or CBF respectively. Eccentricities between 4-6° were combined to represent the signals within the stimuli region. Shaded regions represent the standard error across nine participants.

**Supplemental Figure S6.**
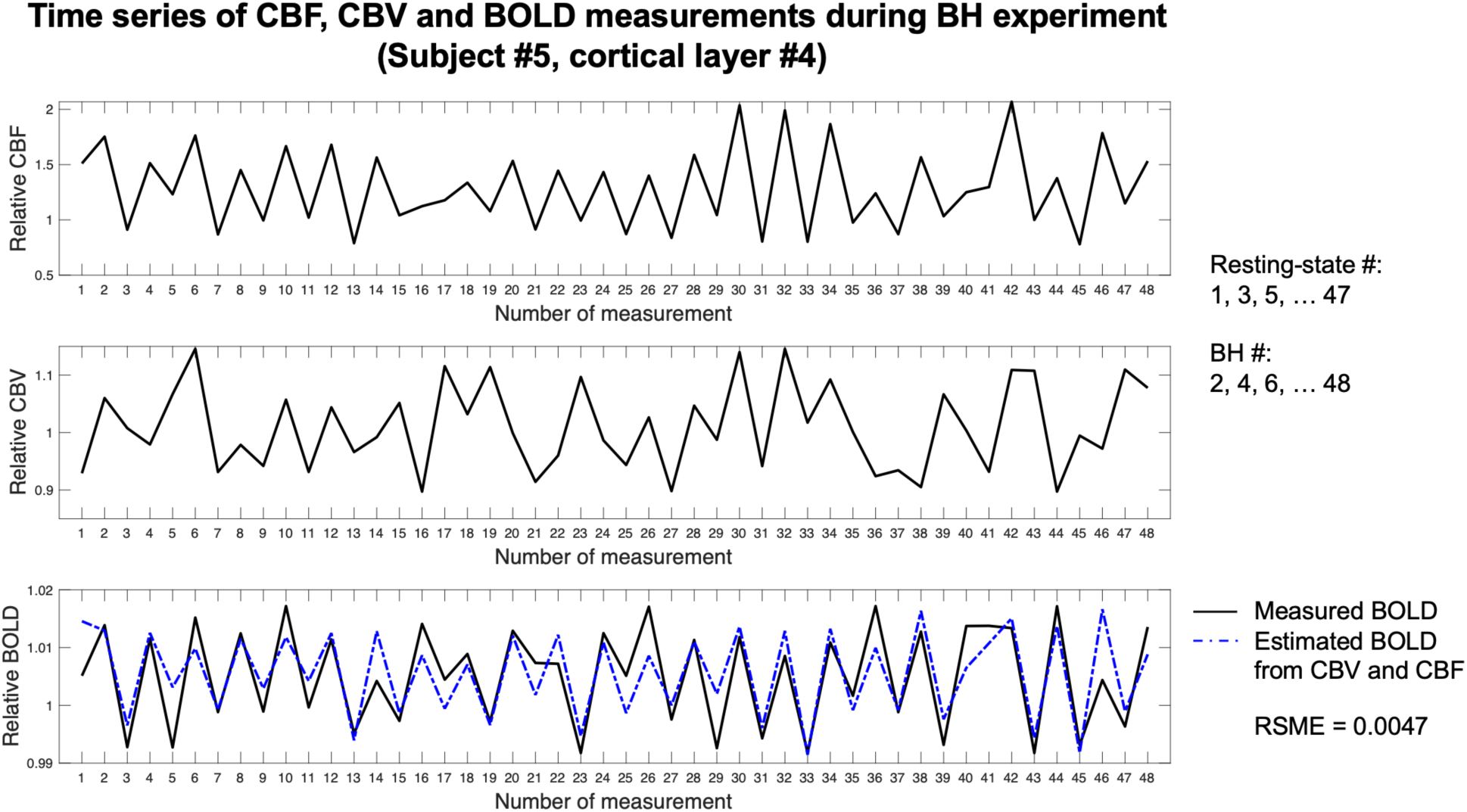
An example of dynamic time series of CBF, CBV, and BOLD signals averaged from cortical layer 4 of participant 5. To normalize the signals, relative CBF, CBV, and BOLD data were divided by their respective averages obtained during resting-state scans (#1, 3, 5, … 47). Increases in CBF, CBV, and BOLD signals during even-numbered measurements are expected due to BH-induced hypercapnia. Variations in the hypercapnia levels during BH and the delayed vascular responses contribute to observed fluctuations throughout the dynamic time series. Consequently, averaging signals from resting-state scans may lead to an overestimation of baseline CBF, CBV, and BOLD levels. The proposed method considers baseline CBF, CBV and BOLD values as unknown parameters in signal fitting, which minimizes the overestimation of baseline signals and improves accuracy for estimation of M and β values. The dashed blue line represents estimated BOLD signals using measured CBF and CBV, along with estimated M and β values according to Eq. [1]. RSME between measured and estimated BOLD signals was 0.0047 for the shown signals, and was 0.0050±0.0019 across all participants and cortical layers.

**Supplemental Figure S7.**
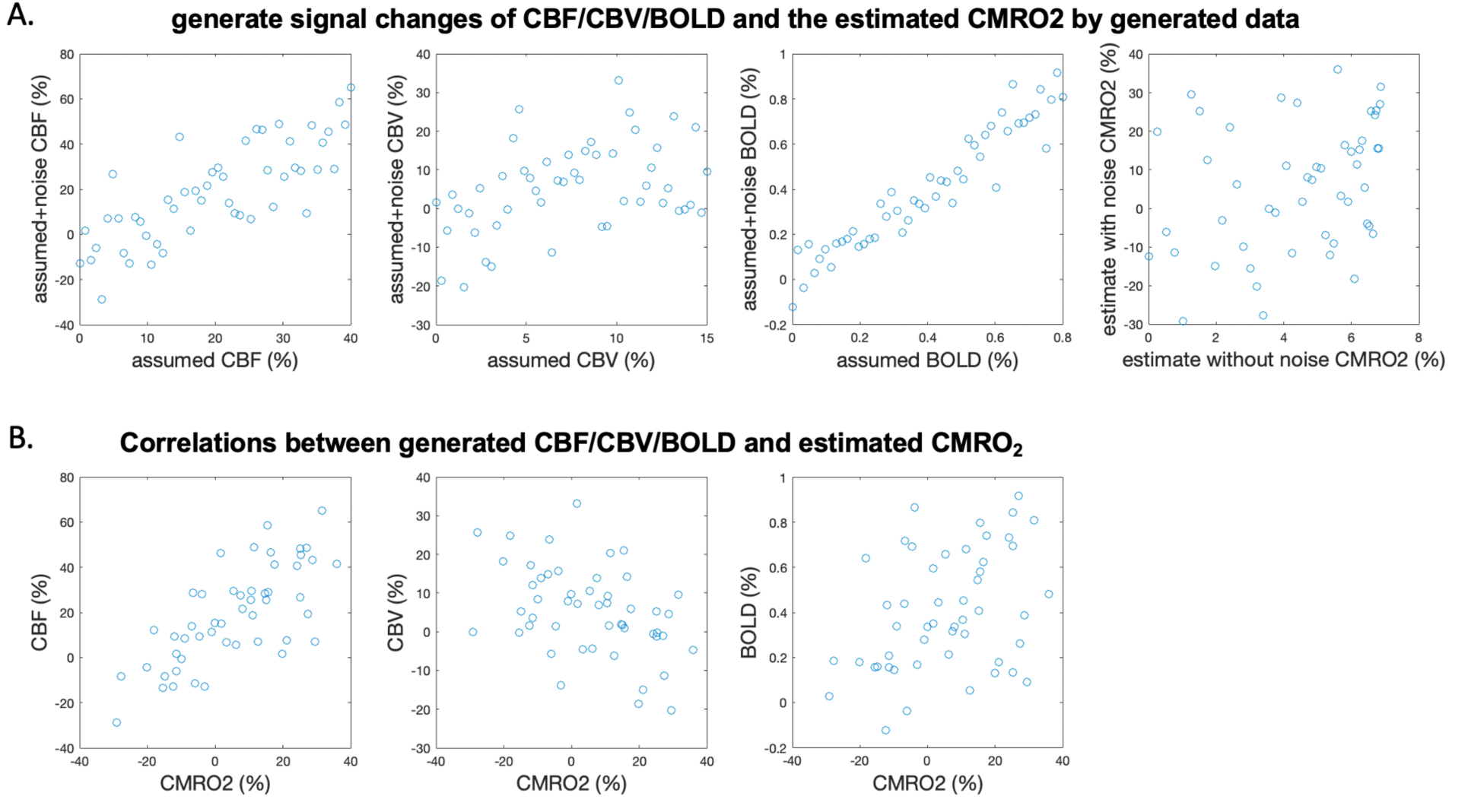
Simulated relationship between CBF/CBV/BOLD and CMRO_2_. (A) Generate signal changes of CBF/CBV/BOLD and the estimated CMRO_2_ by generated data. According to the activation in Fig.7, the CBF/CBV/BOLD signals were simulated. The CBF signal changes from 0-40%, the CBV signal changes from 0-15%, and the BOLD signal changes from 0-0.8%. And M, beta and SNR are generated according to the average value of our data (M = 0.055, beta = 1.1, SNR = 10). Noise is added according to SNR, and the real data is simulated. The first three figures are the relationship between the data generated by CBF/CBV/BOLD and the data after adding noise. The last figure is the relationship between the CMRO_2_ signal estimated by the data without noise and the CMRO_2_ signal estimated by the data with noise. (B) Correlations between generated CBF/CBV/BOLD and estimated CMRO_2_. We find that the results for the simulated data are highly similar to those for the real data, and we can see a relationship that corresponds to the LMM results.

**Supplemental Figure S8.**
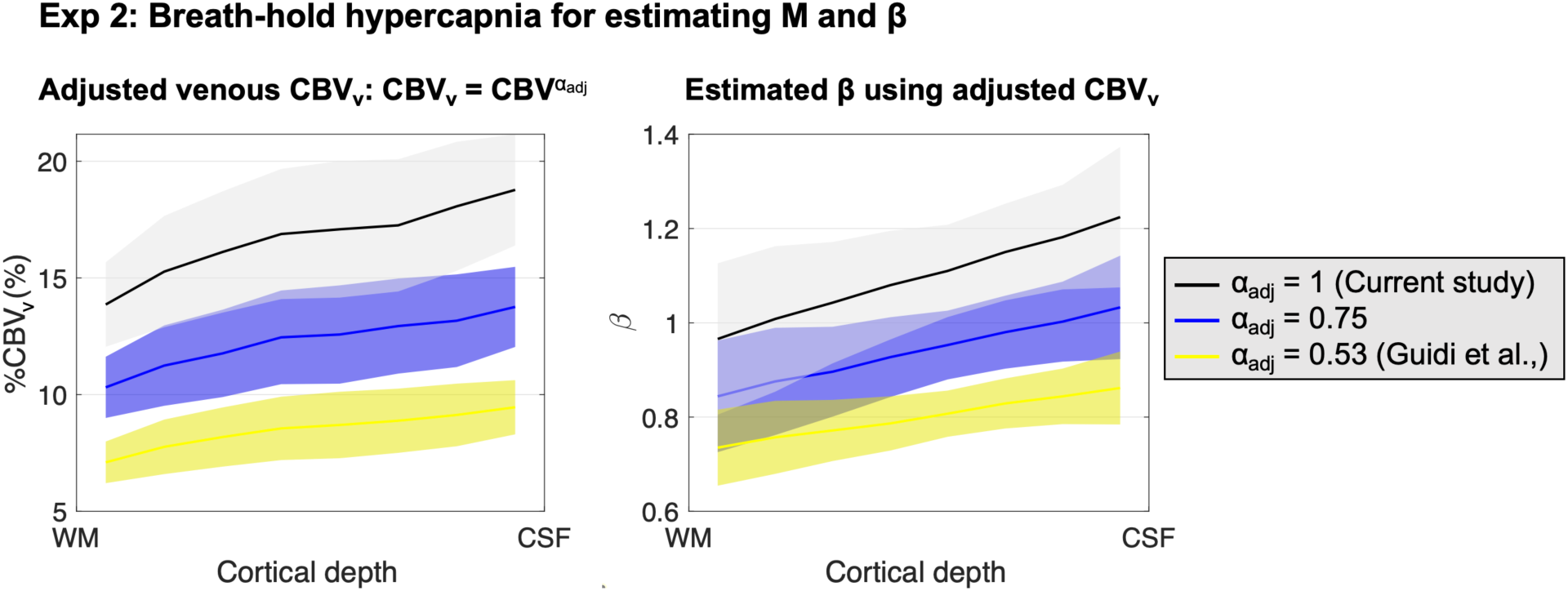
Examples of adjusted venous CBV (CBV_v_ = CBV^αadj^) and corresponding estimated β values across cortical layers in Experient 2 using BH hypercapnia. Examples are shown with α_adj_ = 1 (Current study, CBV_v_ = total CBV), 0.53 (Guidi et al., 2016) and an intermediate value of 0.75.

