## Supplemental for "Laminar multi-contrast fMRI at 7T allows differentiation of neuronal excitation and inhibition underlying positive and negative BOLD responses"

### Supplemental information

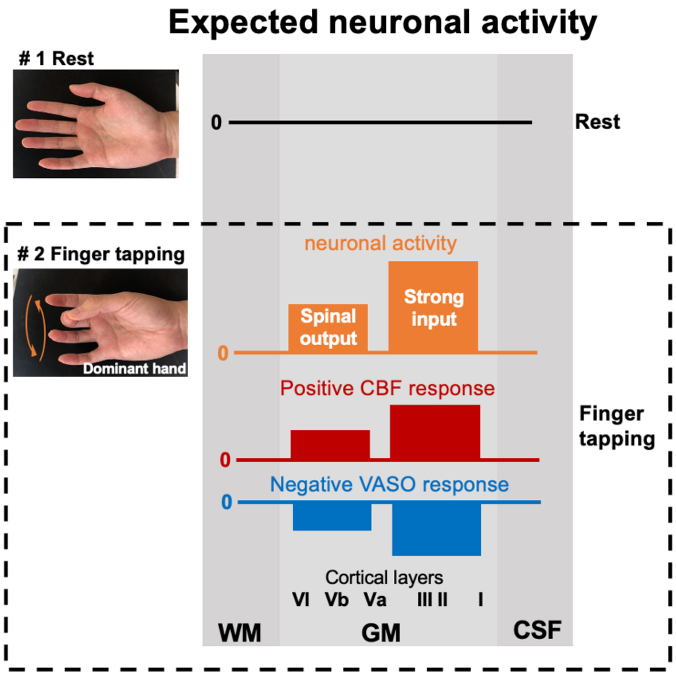


***Supplemental Figure S1: Expected neuronal activation and corresponding CBF and VASO responses to FT.*** *FT is expected to induce neuronal activation across both superficial and deep cortical layers, hypothetically engaging proprioceptive and exteroceptive sensory inputs from the somatosensory (S1) and premotor cortex, and motor output from deep layers. This neuronal activation is expected to associate with double-peak increase in CBF and decrease in VASO (increase in CBV), with stronger responses near the superficial layers.*

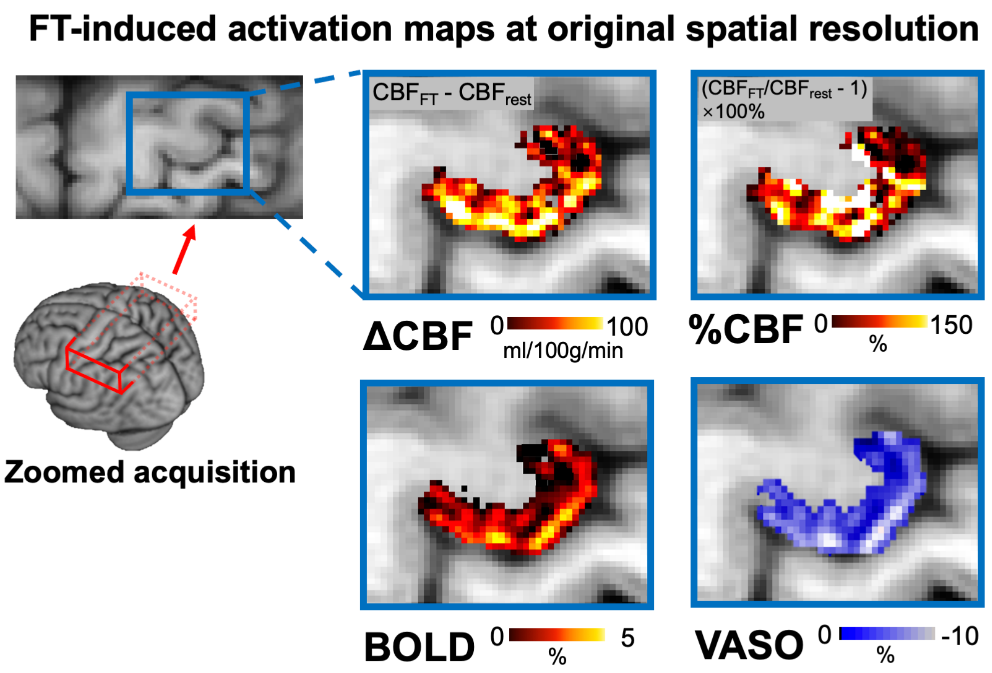


***Supplemental Figure S2. FT induced activation maps at original spatial resolution.*** *These activation maps were shown at the original spatial resolution (1 mm in-plane) before being upsampled to a finer grid for cortical depth analysis. Double-peak activation patterns are still observable at this spatial resolution.*

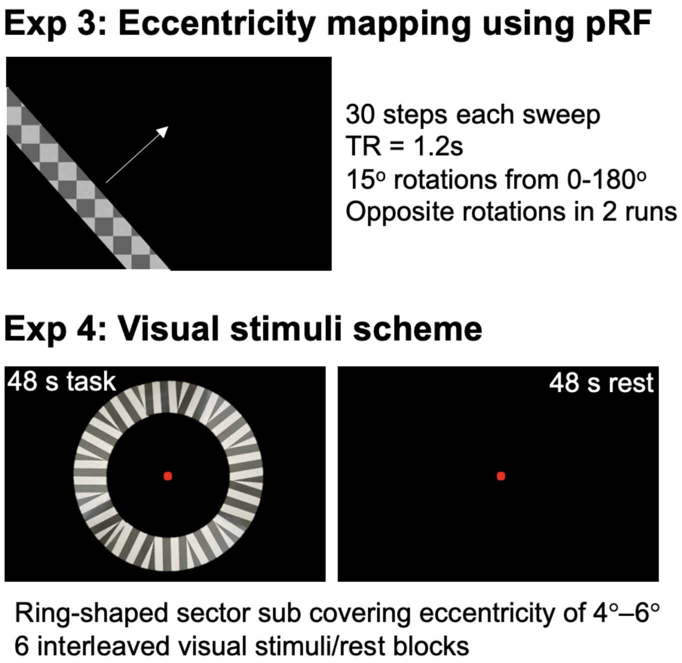


***Supplemental Figure S3. Stimulus schemes for visual stimuli experiments.*** *Top panel shows pRF stimulus pattern for eccentricity mapping (experiment 3): flickering checkerboard pattern that sweeps (30 steps, TR=1.2s) across the screen, sequentially stimulating different visual field regions with a 15-degree rotation after each sweep. The bottom panel shows visual stimulus consisting of a mean-gray background and a ring-shaped sector covering an eccentricity of 4°–6°. Each subsector contained a high-contrast (100%) radial grating (1 cpd) that reversed contrast at 4Hz. One acquisition block (48 sec) acquired two ASL and VASO images and four BOLD images.*

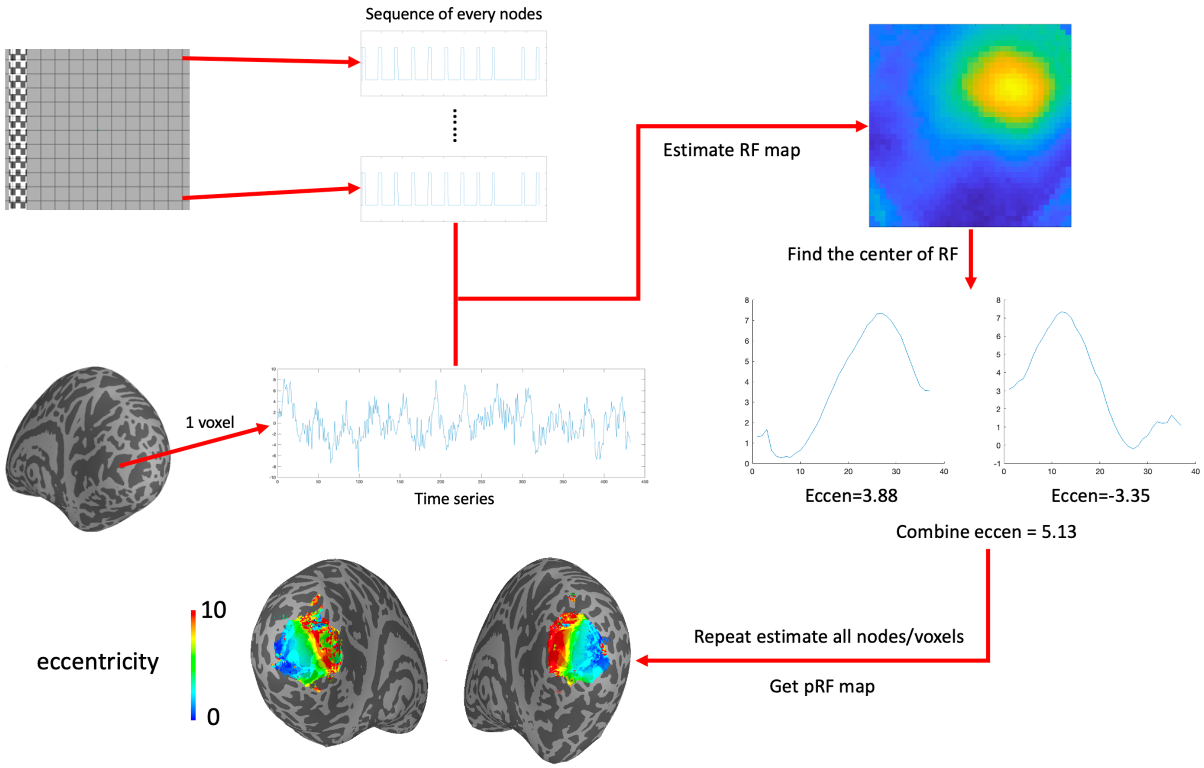


***Supplemental Figure S4. pRF calculation process.*** *First, we sample the visual field into a 40×40 (both the x-axis and y-axis are from -10 degrees to 10 degrees, with a step of 0.5 degrees). As shown in S3, we can obtain the time series of the visual stimulus sweep of each node in the grid. Then we select a voxel to extract its time series. The time series of this voxel is deconvolved and combined with the sequences of all nodes in the grid to calculate the response of the neurons in this voxel to the visual stimulation at each node. In this way we can get the receptive field map of the neurons in this voxel. Then find the center position of the receptive field and calculate its position in the visual field (eccentricity and angle). Then, we can get the centrifugation corresponding to this voxel. We can calculate the eccentricity map by repeating this step for all voxels.*

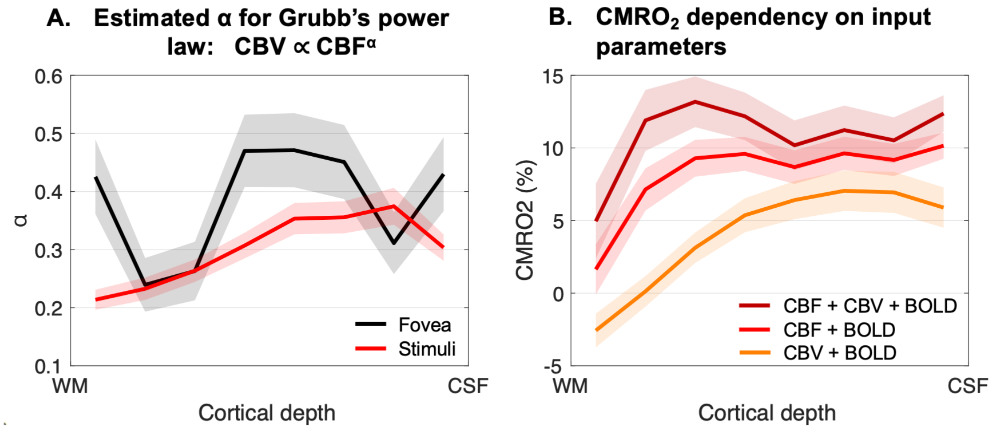


***Supplemental Figure S5.*** *A. Estimated α values in the stimuli and fovea regions using CBF and CBV measurements. B. Comparisons between CMRO2 measured from BOLD and simultaneously measured CBF and CBV (reference, dark red trace), or with only CBF (light red trace) or CBV (orange trace). When only CBF or CBV was used for CMRO2 calculation, the Grubb's power law was utilized to generate the corresponding CBV or CBF respectively. Eccentricities between 4-6° were combined to represent the signals within the stimuli region. Shaded regions represent the standard error across nine participants.*

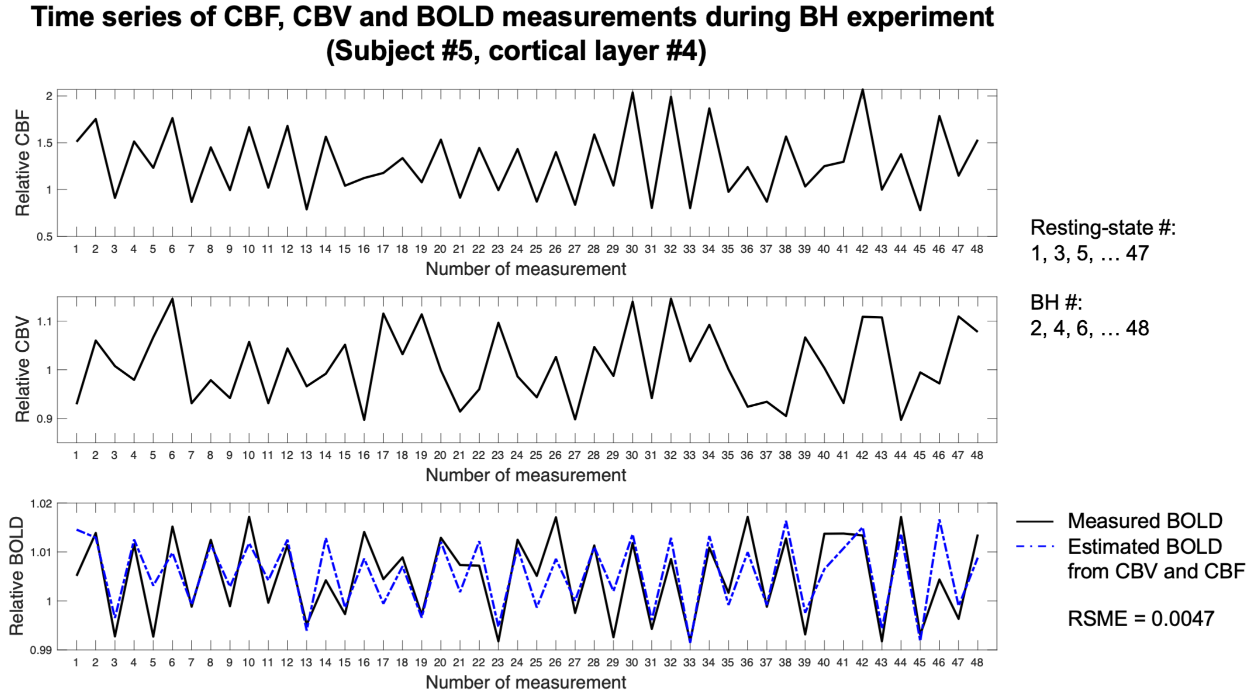


***Supplemental Figure S6.*** *An example of dynamic time series of CBF, CBV, and BOLD signals averaged from cortical layer 4 of participant 5. To normalize the signals, relative CBF, CBV, and BOLD data were divided by their respective averages obtained during resting-state scans (#1, 3, 5, … 47). Increases in CBF, CBV, and BOLD signals during even-numbered measurements are expected due to BH-induced hypercapnia. Variations in the hypercapnia levels during BH and the delayed vascular responses contribute to observed fluctuations throughout the dynamic time series. Consequently, averaging signals from resting-state scans may lead to an overestimation of baseline CBF, CBV, and BOLD levels. The proposed method considers baseline CBF, CBV and BOLD values as unknown parameters in signal fitting, which minimizes the overestimation of baseline signals and improves accuracy for estimation of M and β values. The dashed blue line represents estimated BOLD signals using measured CBF and CBV, along with estimated M and β values according to Eq. [1]. RSME between measured and estimated BOLD signals was 0.0047 for the shown signals, and was 0.0050±0.0019 across all participants and cortical layers.*

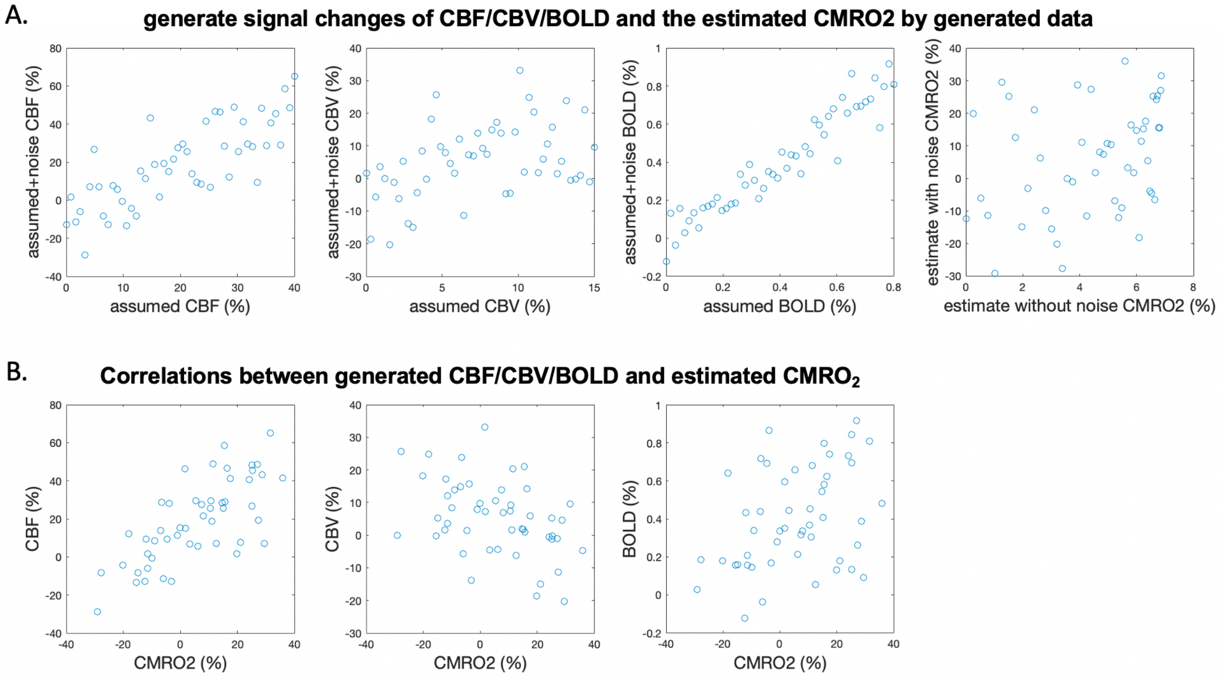


***Supplemental Figure S7.*** *Simulated relationship between CBF/CBV/BOLD and $CMRO_2$. (A) Generate signal changes of CBF/CBV/BOLD and the estimated $CMRO_2$ by generated data. According to the activation in Fig.7, the CBF/CBV/BOLD signals were simulated. The CBF signal changes from 0-40%, the CBV signal changes from 0-15%, and the BOLD signal changes from 0-0.8%. And M, beta and SNR are generated according to the average value of our data (M = 0.055, beta = 1.1, SNR = 10). Noise is added according to SNR, and the real data is simulated. The first three figures are the relationship between the data generated by CBF/CBV/BOLD and the data after adding noise. The last figure is the relationship between the $CMRO_2$ signal estimated by the data without noise and the $CMRO_2$ signal estimated by the data with noise. (B) Correlations between generated CBF/CBV/BOLD and estimated $CMRO_2$. We find that the results for the simulated data are highly similar to those for the real data, and we can see a relationship that corresponds to the LMM results.*

**Exp 2: Breath-hold hypercapnia for estimating M and β**

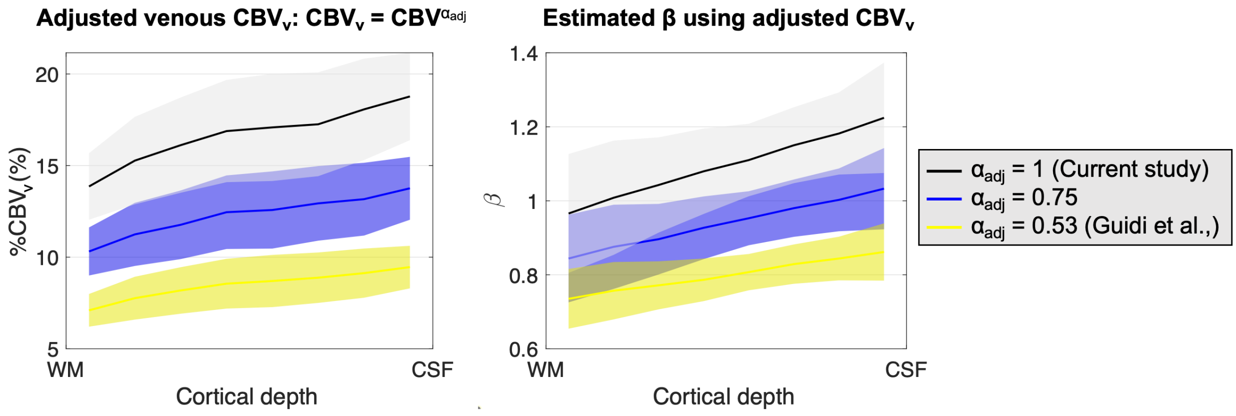


***Supplemental Figure S8.*** *Examples of adjusted venous CBV ($CBV_v = CBV^{\alpha_{adj}}$) and corresponding estimated β values across cortical layers in Experient 2 using BH hypercapnia. Examples are shown with $\alpha_{adj}$ = 1 (Current study, $CBV_v$ = total CBV), 0.53 (Guidi et al., 2016) and an intermediate value of 0.75.*
